# Modelling the impact of extending dose intervals for COVID-19 vaccines in Canada

**DOI:** 10.1101/2021.04.07.21255094

**Authors:** Austin Nam, Raphael Ximenes, Man Wah Yeung, Sharmistha Mishra, Jianhong Wu, Matthew Tunis, Beate Sander

**Author notes:** **Corresponding author:** Austin Nam, Centre for Immunization Readiness, Public Health Agency of Canada, Ottawa, Ontario, Canada.

## Abstract

**Background:** Dual dose SARS-CoV-2 vaccines demonstrate high efficacy and will be critical in public health efforts to mitigate the COVID-19 pandemic and its health consequences; however, many jurisdictions face very constrained vaccine supply. We examined the impacts of extending the interval between two doses of mRNA vaccines in Canada in order to inform deliberations of Canada’s National Advisory Committee on Immunization.

**Methods:** We developed an age-stratified, deterministic, compartmental model of SARS-CoV-2 transmission and disease to reproduce the epidemiologic features of the epidemic in Canada. Simulated vaccination comprised mRNA vaccines with explicit examination of effectiveness against disease (67% [first dose], 94% [second dose]), hospitalization (80% [first dose], 96% [second dose]), and death (85% [first dose], 96% [second dose]) in adults aged 20 years and older. Effectiveness against infection was assumed to be 90% relative to the effectiveness against disease. We used a 6-week mRNA dose interval as our base case (consistent with early program rollout across Canadian and international jurisdictions) and compared extended intervals of 12 weeks, 16 weeks, and 24 weeks. We began vaccinations on January 1, 2021 and simulated a third wave beginning on April 1, 2021.

**Results:** Extending mRNA dose intervals were projected to result in 12.1-18.9% fewer symptomatic cases, 9.5-13.5% fewer hospitalizations, and 7.5-9.7% fewer deaths in the population over a 12-month time horizon. The largest reductions in hospitalizations and deaths were observed in the longest interval of 24 weeks, though benefits were diminishing as intervals extended. Benefits of extended intervals stemmed largely from the ability to accelerate coverage in individuals aged 20-74 years as older individuals were already prioritized for early vaccination. Conditions under which mRNA dose extensions led to worse outcomes included: first-dose effectiveness < 65% against death; or protection following first dose waning to 0% by month three before the scheduled 2^nd^ dose at 24-weeks. Probabilistic simulations from a range of likely vaccine effectiveness values did not result in worse outcomes with extended intervals.

**Conclusion:** Under real-world effectiveness conditions, our results support a strategy of extending mRNA dose intervals across all age groups to minimize symptomatic cases, hospitalizations, and deaths while vaccine supply is constrained.

## Introduction

One year into the coronavirus disease 2019 (COVID-19) pandemic, several vaccines have been approved by regulatory bodies across the globe and recommended by national and international immunization technical advisory groups. Many countries face limited vaccine supplies as manufacturers ramp up capacity while periodically encountering lower production output and interrupted operations. With constrained supply in the early months of 2021, the government of Canada faced questions about how to best allocate available vaccines (which were all two-dose schedules) to meet the public health goal of minimizing serious illness and deaths while minimizing societal disruption as a result of the COVID-19 pandemic. Specifically, should faster vaccine coverage be pursued by extending the time to second dose in exchange for lower of protection with the first dose until the second dose is administered? The question reflects the need to balance individual protection and population impact, given that individuals also benefit from indirect protection when overall SARS-CoV-2 circulation is diminished. That is, an individual’s probability of infection declines faster with higher coverage at a population-level. Canada’s National Advisory Committee on Immunization (NACI) had previously recommended, for Pfizer, Moderna, and AstraZeneca vaccines, that “jurisdictions may consider delaying the second dose due to logistic or epidemiologic reasons until further supplies of the vaccine become available, preferably within 6 weeks of the first dose”.^1^ The Public Health Agency of Canada (PHAC) developed a mathematical model to explore COVID-19 vaccination strategies with longer extended intervals, which were notably being deployed in the province of Quebec and the United Kingdom.^2,3^

Benefits from different vaccination strategies (various extended interval or no delay) may be realized under different conditions such as different dosing intervals, epidemic scenarios, and assumptions about protection against disease, protection against infection and waning. Such data on vaccine performance continue to emerge with new trial data and real-world evidence. The objective of our study was to examine the epidemiological impact of extending dose intervals for vaccination strategies that use mRNA vaccines in the context of constrained vaccine supply in Canada.

## Methods

### Model description

A deterministic compartment model was constructed to represent transmission of SARS-CoV-2 and effects of vaccination on symptomatic disease, hospitalizations, and deaths in the Canadian population excluding residents of long-term care homes. The modelled population was stratified into five-year age groups up to 75+ years, according to 2019 demographic estimates.^4^ Upon acquisition of infection (Figure 1), individuals are initially in a non-infectious latent period after which they either develop asymptomatic infection (infectious) or symptomatic infection (an infectious period preceded by a pre-symptomatic infectious period). Symptomatic individuals may experience mild/moderate or severe disease with the latter receiving hospital care, where they ceased to be infectious.

**Figure 1.**
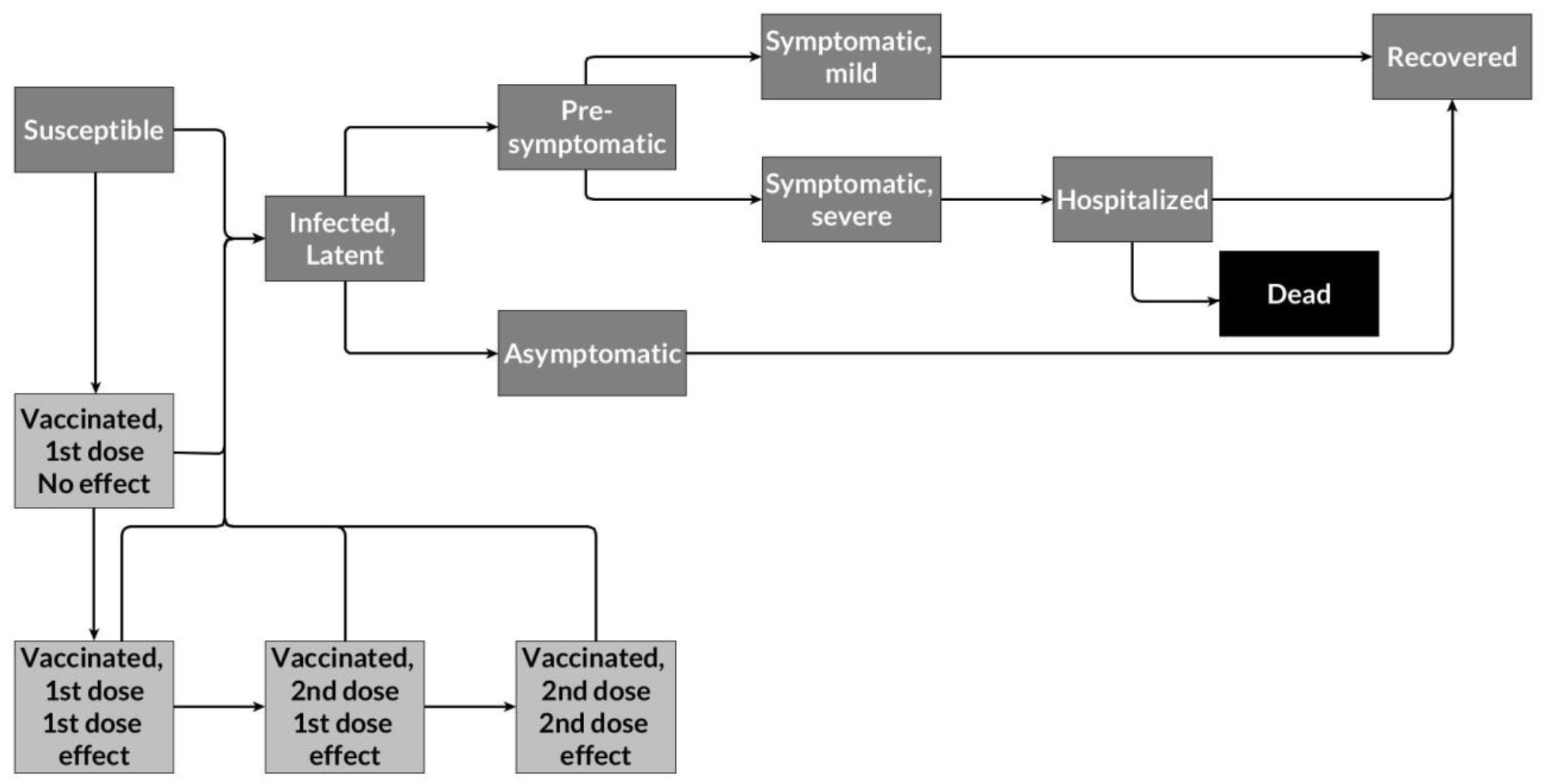
Schematic of general model structure.

Transmission occurred via contacts between susceptible and infectious individuals. Contacts and mixing among age groups were based on the projected daily contacts for Canada.^5^ The impact of public health measures and physical distancing on transmission was modelled as a time-dependent parameter that modulated the force of infection. All asymptomatic and mild/moderate symptomatic infections were assumed to recover while a proportion of individuals with severe symptomatic infections died in hospital. We assumed that individuals with severe symptoms self-isolated until hospitalization and that severe cases did not die without being hospitalized. Transmission from severe symptomatic cases was modelled assuming contact rates in isolation were 25% of the projected home contacts. We assumed severe cases did not contribute to infection once in the hospital setting and did not distinguish between critical cases in intensive care and those in in the hospital ward. We assumed there was no waning immunity following infection-acquired immunity.

Individuals were vaccinated if susceptible or recovered (no previous vaccination). Model assumptions for vaccine performance were established based on available effectiveness studies and consultation with PHAC vaccine experts. Vaccinations were modelled as two-dose regimens with vaccine effectiveness having a joint effect on the force of infection and probabilities of symptomatic disease, hospitalizations, and death. A schematic of the general model structure is shown in Figure 1. The full set of model equations and parameter values are listed in the Supplementary Materials. Model parameters for the transmission model were calibrated by Bayesian inference with prior beliefs and constraints in parameter space informed by literature sources (described in the Supplementary Materials). Key model parameters used in this analysis are listed in Table 1.

**Table 1.** Key model parameters and vaccine characteristics

| Parameter | Value | Source/Rationale |
| --- | --- | --- |
| Transmission coefficient ($\beta$) | 0.0225 | Calibrated |
| Latent period ($1/\rho$), days | 3.96 | Calibrated, informed by Zhao et al.<sup>30</sup> |
| Pre-symptomatic period ($1/\delta_p$), days | 2.48 | Calibrated, informed by He et al.<sup>31</sup> |
| Mild/moderate symptom onset to recovery ($1/\gamma$), days | 5.11 | Calibrated |
| Severe symptom onset to hospitalization ($1/\delta_x$), days | 5.70 | Calibrated, informed by PHAC Weekly Epidemiological Report 17 January to 23 January 2021<sup>32</sup> |
| Length of hospital stay, non-survivors ($\text{losd}_h$) | 19.84 | Calibrated, informed by CIHI<sup>33</sup> |
| Probability of severe/hospitalization \| infected ($\kappa$) | 0-19: 0.0013<br>20-29: 0.0042<br>30-39: 0.0079<br>40-49: 0.0108<br>50-59: 0.0194<br>60-69: 0.0602<br>70-74: 0.1093<br>75+: 0.5298 | Calibrated |
| Probability of death \| hospitalized | 0-19: 0.0064<br>20-29: 0.0180<br>30-39: 0.0614<br>40-49: 0.0767<br>50-59: 0.1880<br>60-69: 0.2273<br>70-74: 0.4055<br>75+: 0.5268 | Calibrated |
| Time to effect, first dose (days) | 14 | Baden et al.<sup>6</sup><br>Polack et al.<sup>7</sup> |
| Time to effect, second dose (days) | 7 | Baden et al.<sup>6</sup><br>Polack et al.<sup>7</sup> |
| Duration of vaccine protection after first dose (years) | 2 | Assumption |

### Vaccination

As of March 12, 2021, three COVID-19 vaccines had been approved for use in Canada: two mRNA vaccines that have demonstrated high efficacy against symptomatic disease and a viral vector vaccine with lower efficacy against symptomatic disease.^6–8^ We considered vaccination programs consisting of an mRNA vaccine, which is the major vaccine in Canada, using different dose intervals (Table 2). In the base case, mRNA vaccines were modelled as a two-dose regimen with a 6-week interval between doses. We then examined the potential impact on symptomatic disease, hospitalizations, and deaths from extending the interval between doses for mRNA vaccines to 12, 16, and 24 weeks. In all scenarios, extended intervals for mRNA vaccines did not begin until March 1, 2021, prior to which a 6-week interval was maintained. Table 2 lists the definitions of the vaccination strategies in this analysis. mRNA vaccines were administered to all adults aged 20 years and older in descending order of age group until age 55 years and then administered proportionally to all individuals aged 20-54 years. We assumed first vaccine coverage of 65% in individuals aged 20-64 years and 80% in ages 65 and older.^9^ Once the vaccine coverage for first dose administration was reached in the age group (65% or 80%), the next prioritized age group would receive their first doses until coverage was reached. All susceptible and recovered individuals were eligible to be vaccinated, but individuals with active infections were not vaccinated. For simplicity, individuals who were infected between the first and second dose did not receive a second dose.

**Table 2.**
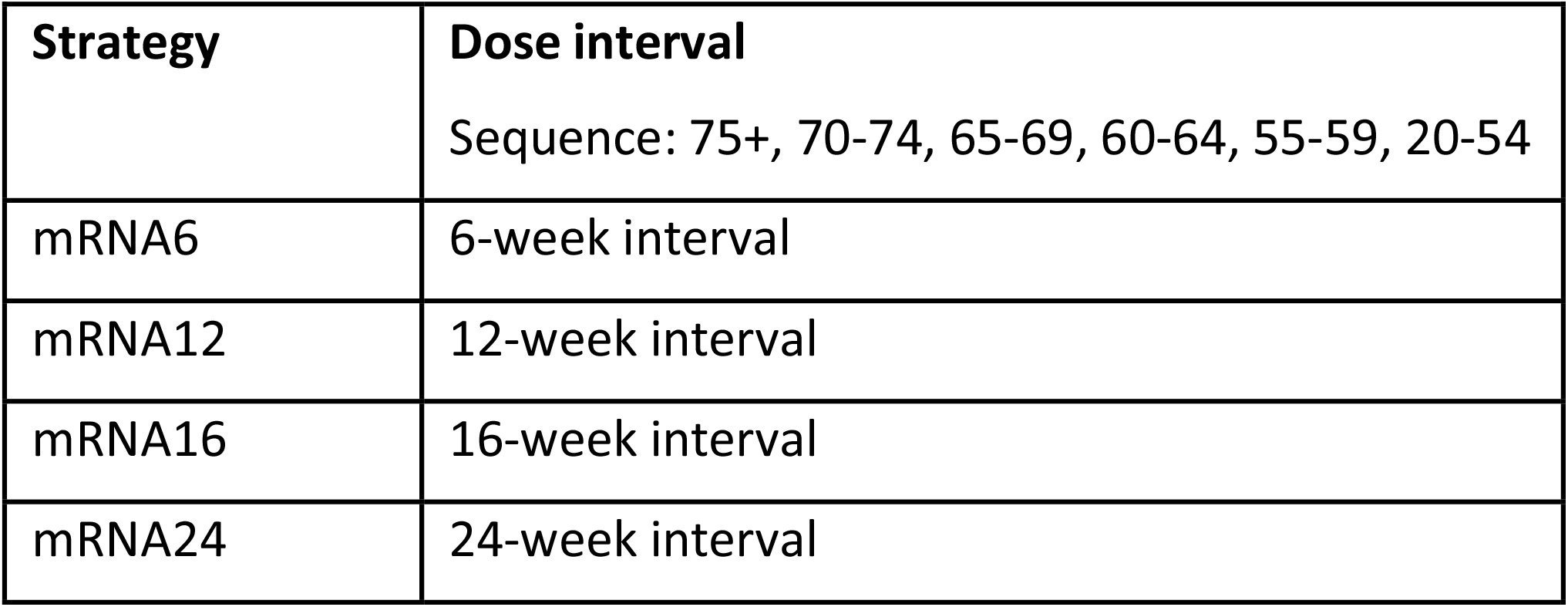
Vaccination strategies using different mRNA dose intervals

The vaccine effectiveness assumptions used in the model are listed in Table 3. We represented vaccine effectiveness as “leaky” protection in which all vaccinated individuals were subject to some residual risk of infection and symptomatic disease. This is in contrast to an “all-or-none” vaccine which confers complete protection to a proportion of vaccinated individuals. We used the same concept of vaccine effectiveness as Swan et al., in which the overall effectiveness against symptomatic disease (VE_dis_) is a function of the risk of infection and risk of symptomatic disease conditional on infection.^10^ Formally, this is represented as, VE_dis_ = 1 - (1 – VE_inf_)(1 – VE_symp_), where VE_inf_ represents effectiveness against infection and VE_symp_ represents the effectiveness against symptomatic disease, conditional on infection. In addition, vaccine effectiveness against hospitalizations was modelled conditional on symptomatic disease (VE_hosp_ = 1 – (1 – VE_dis_)(1 – VE_hosp|disease_) and vaccine effectiveness against deaths was modelled conditional on hospitalizations (VE_death_ = 1 – (1 – VE_hosp_)(1 – VE_death|hosp_). We assumed that vaccine effectiveness against infections was 90% of the effectiveness against symptomatic disease. Recent real-world effectiveness estimates suggest that mRNA vaccines may be almost as effective at preventing infections as they are at preventing symptomatic disease.^11–16^ First-dose effectiveness values (Table 3) were based on estimates from the United Kingdom, where an extended dose interval strategy was employed.^13,16–19^ Second-dose effectiveness values (Table 3) were based on estimates from Israel, where >50% of the population had received two doses by early March.^20,21^ For both vaccines, protection began 14 and 7 days after administrating the first and second doses, respectively.^6,7^ We used a mild waning effect in our base case analysis with an average duration of protection of two years under a single dose (protection dropped to 0% in 2 years or protected individuals became susceptible at a rate of 1% per week) and interrogated the impact of waning protection by examining durations of protection of 3-6 months in sensitivity analysis. We did not examine waning protection after the second dose due to the short time horizon used for this analysis (12 months). We also tested sensitivity of our results to first dose VE_hosp_ and VE_death_ values between 50% and 85% while holding second dose effectiveness at their base case values.

**Table 3.** Vaccine effectiveness assumptions

| **Vaccine effectiveness** | **Base case value (range for probabilistic analysis)** | **Distribution (for probabilistic analysis)** |
| --- | --- | --- |
| VE Infection | 90% x VE Disease (80-95%) | Uniform (0.8, 0.95) |
| VE Infection (conservative) | 50% x VE Disease (40-60%) | Uniform (0.4, 0.6) |
| VE Disease<br>Dose 1, <65 years | 67% (48-79%)<sup>13</sup> | Beta (22.18, 11.56) |
| VE Disease<br>Dose 1, 65+ years | 58% (36-71%)<sup>17</sup> | Beta (16.28, 12.79) |
| VE Disease<br>Dose 2, 20+ years | 94% (87-98%)<sup>20,21</sup> | Beta (63.27, 4.13) |
| VE Hospitalization<br>Dose 1, 20+ years | 80% (70-85%)<sup>16,17,19</sup> | Beta (84.85, 22.52) |
| VE Hospitalization<br>Dose 2, 20+ years | 96% (95-97%)<sup>20</sup> | Beta (2888.52, 121.46) |
| VE Death<br>Dose 1, 20+ years | 85% (75-92%)<sup>17,18</sup> | Beta (55.59, 9.99) |
| VE Death<br>Dose 2, 20+ years | Assumed same value as Dose 2 VE Hospitalization |  |

We examined the joint uncertainty in our vaccine effectiveness assumptions by running probabilistic simulations using 2,000 samples. The distributions used to draw random values represented our current belief about the range and distribution of likely effectiveness values (Table 3). We also ran probabilistic simulations to examine a conservative scenario of low effectiveness against infection.

Vaccine uptake was constrained by the available vaccine supply and the capacity to administer doses. The explicit supply schedule used in this analysis is provided in the Supplementary Materials and was based on public announcements.^22,23^ We assumed that half the weekly supply would be reserved for the second dose with a 6-week interval^24^ but extended intervals would allow for the entire weekly supply to be used. The maximum daily rate of administration was assumed to be 150,000 doses in January-March 2021 and increased to 350,000 in April 2021, 450,000 in May 2021, and 525,000 in June-December 2021. The rate of vaccination was explicitly constrained to be the lesser of the total number of doses available and the maximum daily rate of vaccination. We assumed that individuals eligible for their second dose would take priority over those waiting to receive their first dose.

### Model scenarios

We calibrated our model to four calibration targets using data from Ontario, Canada, up to December 18, 2020: daily hospital admissions, daily deaths (excluding long-term care), cumulative hospitalizations by age, and cumulative deaths by age (excluding long-term care). We defined an epidemic trajectory (in the absence of vaccinations) of decreasing infections (R_eff_ = 0.9) starting December 18, 2020 and simulated a third wave beginning on April 1, 2021 (R_eff_ = 1.2), using a time-dependent parameter representing the aggregate effect of different levels of public health measures and physical and social distancing on the force of infection. We did not simulate any additional interventions after the simulated third wave (i.e. modulate the time-dependent parameter) to change the epidemic trajectory (other than vaccinations). We then simulated vaccinations beginning on January 1, 2021 until January 1, 2022 and examined the impact of extending dose intervals for mRNA vaccines starting on March 1, 2021. We also simulated third waves of varying severity (R_eff_ = 1.1, R_eff_ = 1.3) to examine the role of different epidemic scenarios on our assumptions.

## Results

### Vaccine uptake

Figure 2 shows the progression of cumulative vaccinations with the first and second dose for the different vaccination strategies. Extension of the dose interval for mRNA vaccines resulted in accelerated coverage with the first dose in individuals aged 20-74 years with longer intervals having greater impact on younger individuals due to prioritization by age (Figure 3). Under the supply and rollout scenarios used in this model, extending the interval for mRNA vaccines from 6 weeks to 24 weeks advanced the time to coverage for all individuals aged 20 years and older by 33 days, from August 4, 2021 to July 2, 2021.

**Figure 2.**
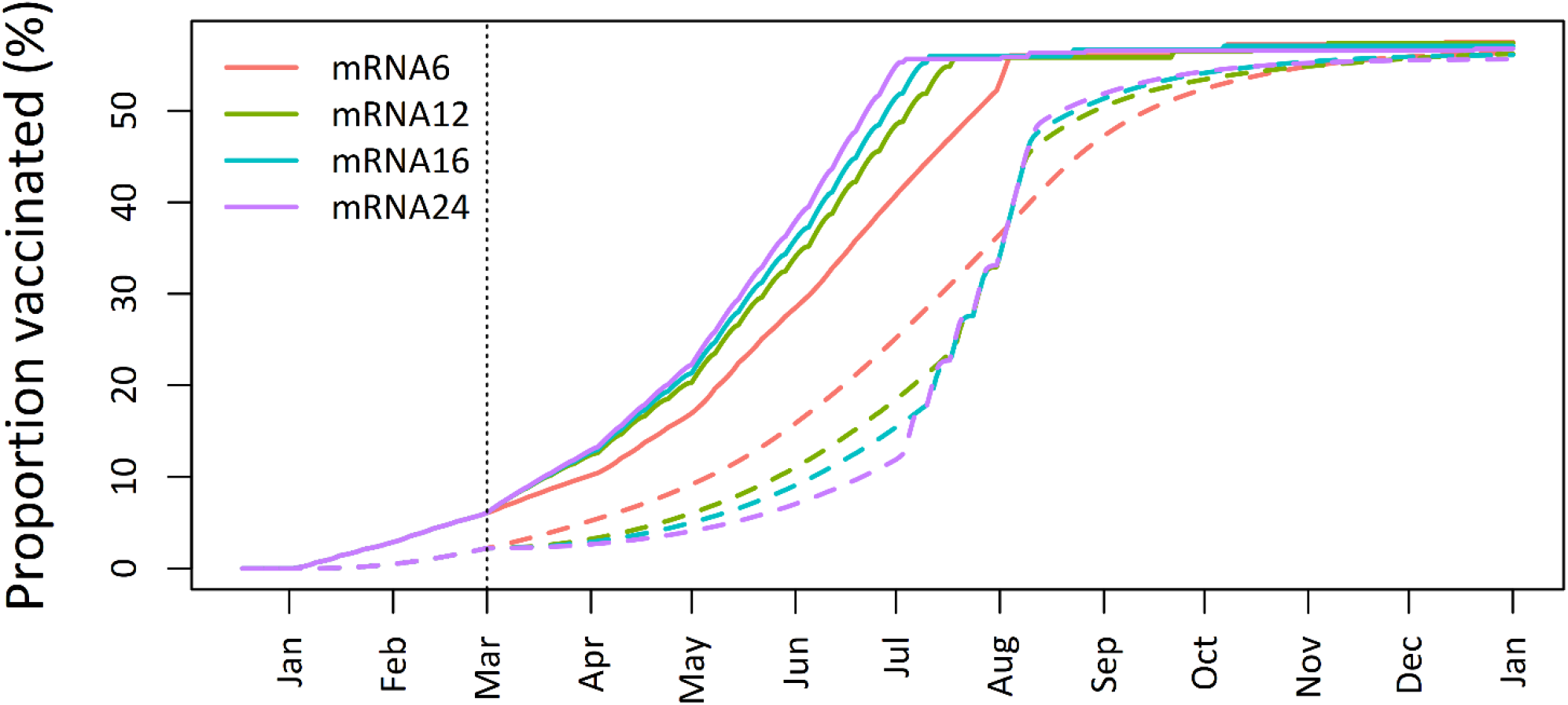
Cumulative vaccinations. Dashed lines represent second dose. Vertical dotted line represents beginning of extended intervals for mRNA vaccines.

**Figure 3.**
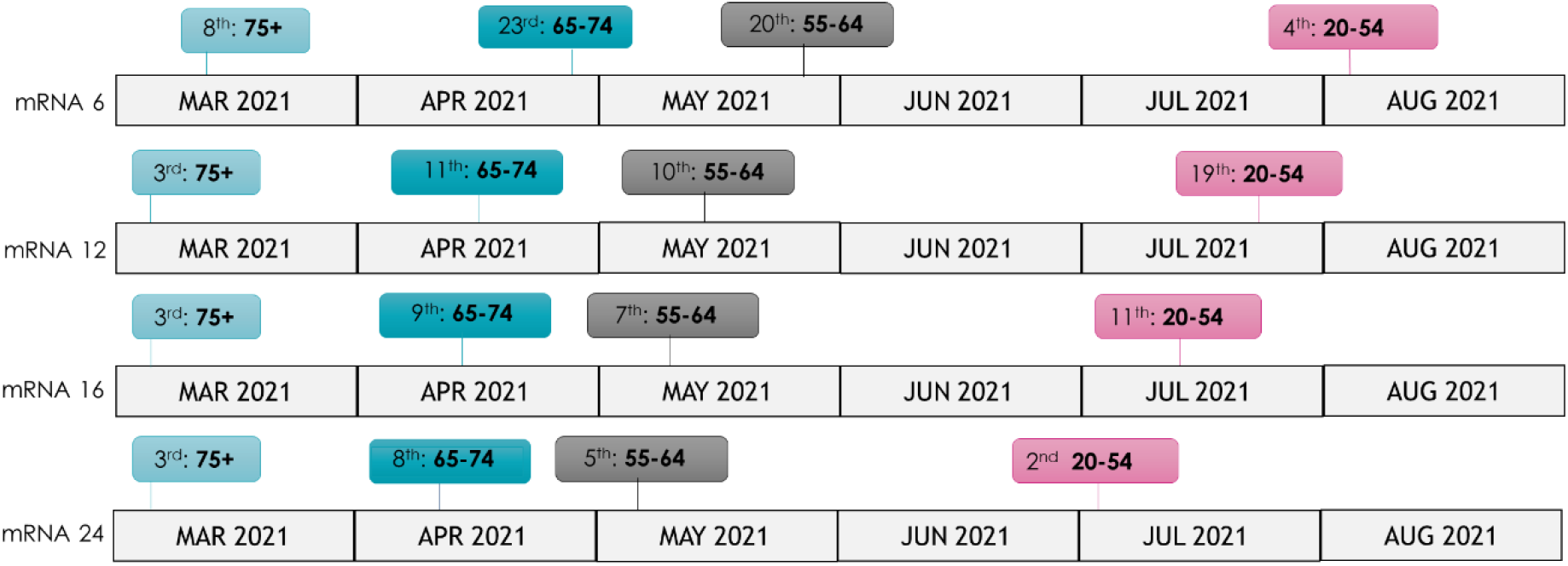
Time to coverage (65% in 20-64 years and 80% in 65+ years) with first dose.

### Population impact of extended dose intervals

Figure 4 and Table 4 show the cumulative incidence of symptomatic disease, hospitalizations, and deaths projected under different vaccination strategies. Under a base case mRNA dose interval of 6 weeks, the model projected cumulative incidence of symptomatic disease, hospitalizations, and deaths in the population of 5,387, 76.09, and 15.53 per 100,000 12 months after the start of the vaccination campaign. Compared to a 6 week interval for mRNA vaccines, dose intervals of 12 to 24 weeks resulted in 12.1-18.9% fewer cases of symptomatic disease (651-1,020 per 100,000), 9.5-13.5% (7.23-10.27 per 100,000) fewer hospitalizations, and 7.5-9.7% (1.16-1.51 per 100,000) fewer deaths. Figure 5 shows the reductions in symptomatic disease, hospitalizations, and deaths of extended intervals at 12 months compared to a 6-week interval from probabilistic simulations. Over the range of sampled values, symptomatic disease, hospitalizations, and deaths decreased in the population with longer intervals. None of the sampled scenarios resulted in worse outcomes compared to a 6-week interval.

**Table 4.** Cumulative incidence of symptomatic disease, hospitalizations, and deaths per 100,000 at 12 months when VE_inf_ = 90% VE_dis_. Relative reductions in parentheses.

| Program Name | Symptomatic disease | Hospitalizations | Deaths |
| --- | --- | --- | --- |
| mRNA6 | 5,387 (Ref) | 76.09 (Ref) | 15.53 (Ref) |
| mRNA12 | 4,736 (12.1%) | 68.86 (9.5%) | 14.37 (7.5%) |
| mRNA16 | 4,571 (15.2%) | 67.34 (11.5%) | 14.15 (8.9%) |
| mRNA24 | 4,368 (18.9%) | 65.82 (13.5%) | 14.02 (9.7%) |

**Figure 4.**
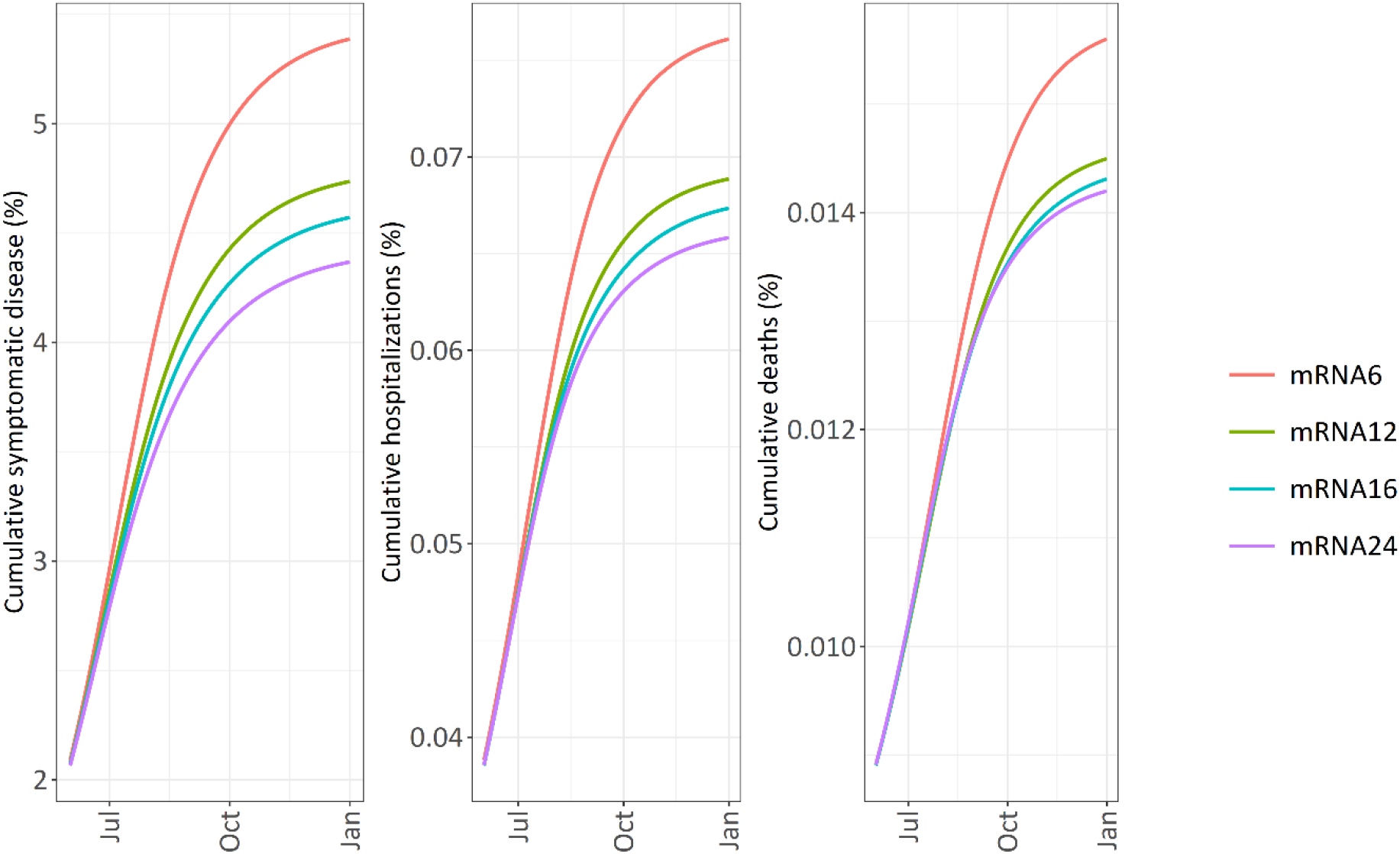
Cumulative incidence of symptomatic disease, hospitalizations, and deaths starting six months after beginning of vaccination campaign.

**Figure 5.**
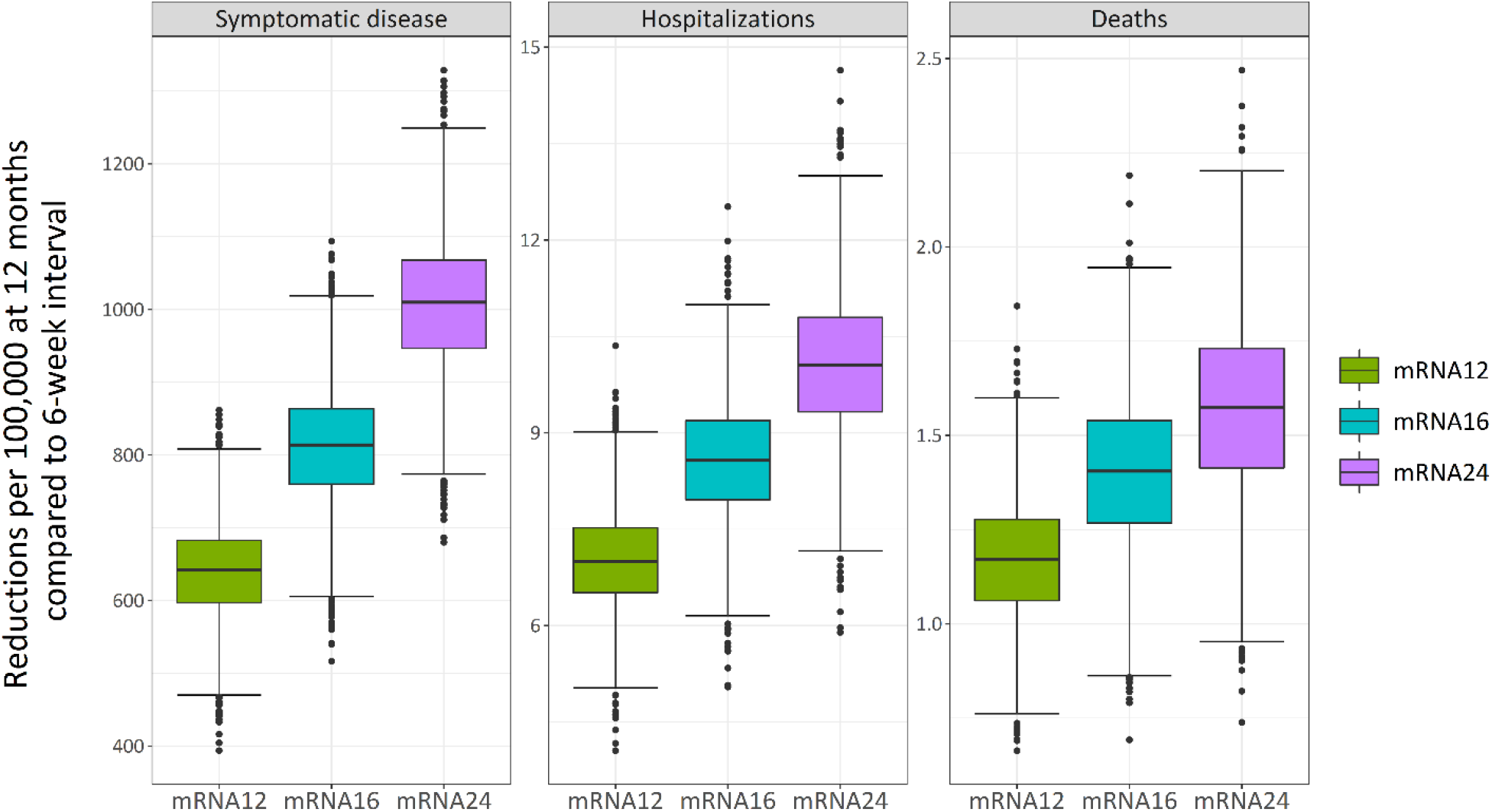
Reductions in symptomatic disease, hospitalizations, and deaths at 12 months compared to a 6-week interval (mRNA6) from probabilistic simulations of 2,000 samples. VE_inf_ = 80-95% VE_dis_.

In a scenario where the vaccines offered lower protection against infection (VE_inf_ = 50% VE_dis_), a 6-week mRNA dose interval was projected to result in cumulative incidence of symptomatic disease, hospitalizations, and deaths in the population of 6,931, 94.39, and 18.68 per 100,000 at 12 months. Extending the mRNA dose interval resulted in 9.2-14.9% (639-1,030 per 100,000) fewer cases of symptomatic disease, 7.6-10.8% (7.17-10.2 per 100,000) fewer hospitalizations, and 6.3-8.1% (1.18-1.51 per 100,000) fewer deaths (Table 5). Probabilistic simulations of vaccine effectiveness values assuming a lower VE_inf_ (40-60% of VE_dis_) showed extended intervals reduced symptomatic disease, hospitalizations, and deaths at 12 months compared to a 6-week interval (Supplementary Materials). None of the sampled values resulted in worse outcomes.

**Table 5.** Cumulative incidence of symptomatic disease, hospitalizations, and deaths per 100,000 at 12 months when VE_inf_=50% VE_dis_. Relative reductions in parentheses.

| Program Name | Symptomatic disease | Hospitalizations | Deaths |
| --- | --- | --- | --- |
| mRNA6 | 6,931 (Ref) | 162.01 (Ref) | 33.93 (Ref) |
| mRNA12 | 6,292 (9.2%) | 87.22 (7.6%) | 17.50 (6.3%) |
| mRNA16 | 6,095 (12.1%) | 85.27 (9.7%) | 17.23 (7.8%) |
| mRNA24 | 5,902 (14.9%) | 84.19 (10.8%) | 17.17 (8.1%) |

### Subgroup analysis

Tables 6 and 7 show the cumulative incidence of symptomatic disease, hospitalizations, and deaths per 100,000 at 12 months by age group. The model projected the largest reductions in hospitalizations and deaths with a 24-week interval for individuals aged 20-74 and with a 16-week interval for individuals aged 75+ years. Longer intervals (16 or 24 weeks) were not optimal in reducing the less critical outcome of symptomatic disease, and a 24-week interval resulted in an increase in symptomatic disease in individuals aged 75+ years compared to a 6-week interval. A scenario of lower effectiveness against infection (VE_inf_ = 50% VE_dis_) projected similar benefits of extended intervals but showed an increase in symptomatic disease in individuals aged 65-74 years compared to a 6-week interval. As all intervals reduced hospitalizations (i.e. severe infections) in the model, increases in symptomatic disease were mild/moderate cases.

**Table 6.** Cumulative incidence of symptomatic disease, hospitalizations, and deaths per 100,000 by age group at 12 months when VE_inf_=90% VE_dis_. Relative reductions in parentheses. Bolded cells indicate the strategy with the largest reductions in outcomes for each age group.

| Age group | Strategy | Symptomatic disease | Hospitalizations | Deaths |
| --- | --- | --- | --- | --- |
| 75+ | mRNA6 | 543.25 (Ref) | 276.97 (Ref) | 117.05 (Ref) |
|  | mRNA12 | **538.32 (9.1%)** | 263.79 (4.8%) | 110.30 (5.8%) |
|  | mRNA16 | 546.46 (5.9%) | **262.72 (5.1%)** | **109.05 (6.8%)** |
|  | mRNA24 | 559.11 (-2.9%)* | 265.42 (4.2%) | 109.44 (6.5%) |
| 65-74 | mRNA6 | 1,476.92 (Ref) | 115.24 (Ref) | 26.90 (Ref) |
|  | mRNA12 | **1,438.94 (2.6%)** | 107.16 (7.0%) | 24.73 (8.1%) |
|  | mRNA16 | 1,447.70 (2.0%) | 106.86 (7.3%) | 24.71 (8.1%) |
|  | mRNA24 | 1,446.46 (2.1%) | **105.26 (8.7%)** | **24.19 (10.1%)** |
| 55-64 | mRNA6 | 2,961.92 (Ref) | 99.33 (Ref) | 13.54 (Ref) |
|  | mRNA12 | 2,693.64 (9.1%) | 89.37 (10.0%) | 12.17 (10.1%) |
|  | mRNA16 | 2,617.86 (11.6%) | 86.36 (13.1%) | 11.75 (13.2%) |
|  | mRNA24 | **2,541.40 (14.2%)** | **83.76 (15.7%)** | **11.40 (15.8%)** |
| 40-54 | mRNA6 | 6,030.52 (Ref) | 82.99 (Ref) | 6.69 (Ref) |
|  | mRNA12 | 5,180.47 (14.1%) | 71.12 (14.3%) | 5.76 (13.9%) |
|  | mRNA16 | 4,981.75 (17.4%) | 68.30 (17.7%) | 5.54 (17.2%) |
|  | mRNA24 | **4,712.7 (21.9%)** | **64.69 (22.1%)** | **5.26 (21.4%)** |
| 20-39 | mRNA6 | 5,839.58 (Ref) | 34.52 (Ref) | 0.91 (Ref) |
|  | mRNA12 | 5,030.73 (13.9%) | 29.61 (14.2%) | 0.78 (14.3%) |
|  | mRNA16 | 4,817.54 (17.5%) | 28.34 (17.9%) | 0.75 (17.6%) |
|  | mRNA24 | **4,568.67 (21.8%)** | **26.87 (22.2%)** | **0.71 (22.0%)** |

\*Increase in cumulative incidence at 12 months compared to mRNA6.

**Table 7.** Cumulative incidence of symptomatic disease, hospitalizations, and deaths per 100,000 by age group at 12 months when VE_inf_=50% VE_dis_. Relative reductions in parentheses. Bolded cells indicate the strategy with the largest reductions in outcomes for each age group.

| Age group | Strategy | Symptomatic disease | Hospitalizations | Deaths |
| --- | --- | --- | --- | --- |
| 75+ | mRNA6 | 682.81 (Ref) | 341.68 (Ref) | 140.62 (Ref) |
|  | mRNA12 | **673.43 (1.4%)** | 325.55 (4.7%) | 132.68 (5.6%) |
|  | mRNA16 | 680.13 (0.4%) | **323.23 (5.4%)** | **130.87 (6.9%)** |
|  | mRNA24 | 702.46 (-2.9%)* | 330.22 (3.4%) | 132.33 (5.9%) |
| 65-74 | mRNA6 | 1,804.03 (Ref) | 137.27 (Ref) | 31.19 (Ref) |
|  | mRNA12 | 1,819.96 (-0.9%)* | 133.80 (2.5%) | 30.03 (3.7%) |
|  | mRNA16 | **1,815.42 (-0.6%)\*** | 132.91 (3.2%) | 30.09 (3.5%) |
|  | mRNA24 | 1,844.66 (-2.2%)* | **132.33 (3.6%)** | **29.47 (5.5%)** |
| 55-64 | mRNA6 | 3,833.59 (Ref) | 127.87 (Ref) | 17.08 (Ref) |
|  | mRNA12 | 3,568.23 (6.9%) | 117.44 (8.2%) | 15.64 (8.4%) |
|  | mRNA16 | 3,483.86 (9.1%) | 114.02 (10.8%) | 15.15 (11.3%) |
|  | mRNA24 | **3,421.87 (10.7%)** | **111.64 (12.7%)** | **14.82 (13.2%)** |
| 40-54 | mRNA6 | 7,532.01 (Ref) | 102.25 (Ref) | 8.11 (Ref) |
|  | mRNA12 | 6,667.78 (11.5%) | 90.05 (11.9%) | 7.15 (11.8%) |
|  | mRNA16 | 6,417.77 (14.8%) | 86.59 (15.3%) | 6.88 (15.2%) |
|  | mRNA24 | **6,130.28 (18.6%)** | **82.75 (19.1%)** | **6.60 (18.6%)** |
| 20-39 | mRNA6 | 7,338.11 (Ref) | 42.81 (Ref) | 1.11 (Ref) |
|  | mRNA12 | 6,492.86 (11.5%) | 37.65 (12.1%) | 0.98 (11.7%) |
|  | mRNA16 | 6,226.69 (15.1%) | 36.10 (15.7%) | 0.94 (15.3%) |
|  | mRNA24 | **5,983.07 (18.5%)** | **34.60 (19.2%)** | **0.90 (18.9%)** |

\*Increase in cumulative incidence at 12 months compared to mRNA6.

### Role of dose 1 effectiveness

Figure 6 shows the cumulative incidence of hospitalizations and deaths over a range of dose 1 VE_hosp_ and VE_death_ values, while VE_dis_ was held at 50% and dose 2 VE_hosp_ and VE_death_ were held at their base case values. Extending the dose interval was projected to reduce hospitalizations at VE_hosp_ values as low as 50% though benefits between a 12-week and 24-week interval became imperceptible at dose 1 VE_hosp_ of 50%. At dose 1 VE_death_ of 65% and 70%, extending the dose interval to 12-24 weeks was projected to increase deaths in the period up until approximately October 2021. However, all extended intervals decreased overall deaths by January 2022 at dose 1 VE_death_ of at least 65%. At a dose 1 VE_hosp_ less than 65%, 16-week and 24-week intervals resulted in an increase in overall deaths. Examination of additional third wave scenarios showed that extended intervals would reduce deaths at lower VE_death_ values if the third wave was more severe than the base case (Supplementary Materials). Conversely, if the third wave was less severe than the base case higher VE_death_ values were needed to reduce deaths.

**Figure 6.**
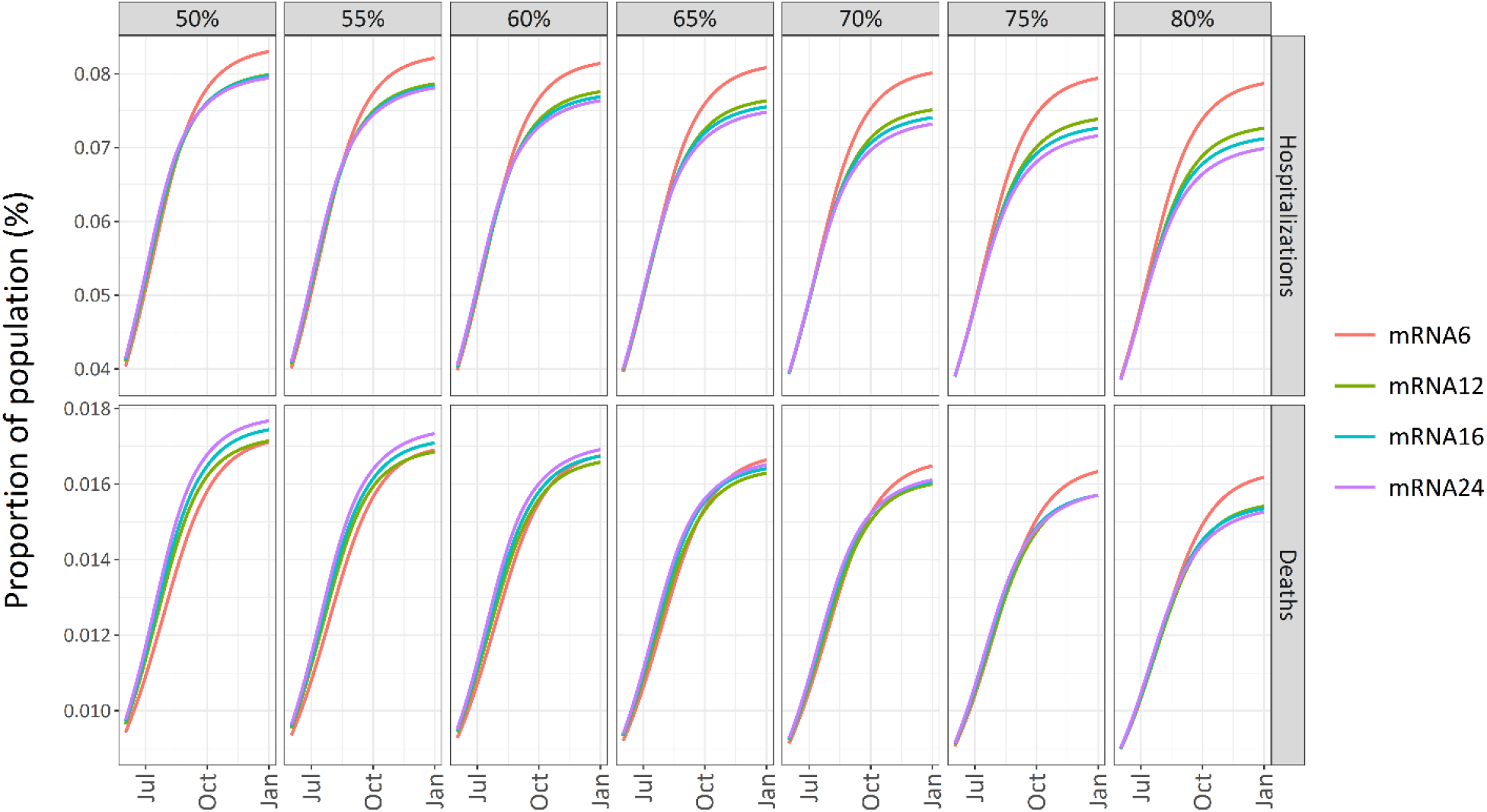
Sensitivity analysis: cumulative incidence of hospitalizations and deaths over different dose 1 VE_hosp_ and VE_death_ values starting six months after the beginning of the vaccination campaign. VE_inf_ = 90% VE_dis_ and VE_dis_ = 50%.

### Role of dose 1 duration of protection

Figure 7 shows the cumulative incidence of symptomatic disease, hospitalizations, and deaths over a range of dose 1 durations of protection (three to six months). In all scenarios, extension of the dose interval reduced overall symptomatic disease and hospitalizations. In a scenario where the average duration of protection after dose 1 was three months, the model projected a small increase in deaths with a 24-week interval.

**Figure 7.**
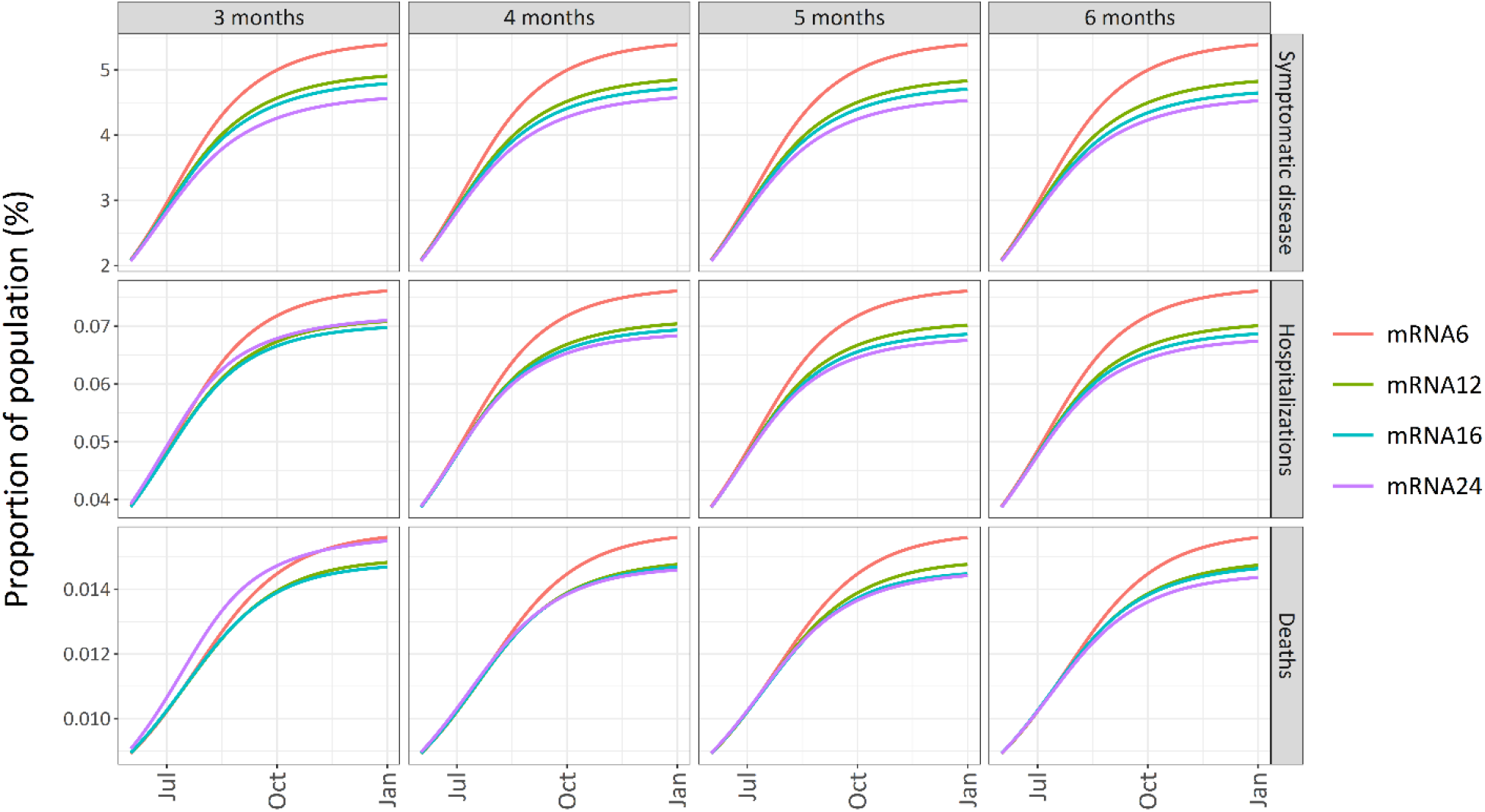
Sensitivity analysis: cumulative incidence of symptomatic disease, hospitalizations, and deaths over different durations of protection after dose 1.

## Discussion

Our model projected that longer mRNA dose intervals (between 12 and 24 weeks) would increase public health benefits in terms of fewer symptomatic cases, hospitalizations, and deaths while vaccine supply is constrained. Overall, our findings show that extending the dose interval conferred benefits to the population by accelerating coverage in individuals lower in the prioritization sequence (Figure 3). Concordant with this, our model also projected a diminishing rate of return in preventing serious outcomes as the dose interval became longer but higher risk age groups were already vaccinated with shorter intervals. It is important to note that these findings are presented in the context of an assumed third wave beginning in April 2021. If a third wave can be avoided or delayed, then the benefits of extending the dose interval would likely diminish.

Two conditions led to worse outcomes (increased deaths) with extending the mRNA dose interval. The first resulted from an average dose 1 duration of protection of three months (i.e. protection dropped to 0% in three months). At the time of this study, we were unaware of any indications that protection from the first dose is waning at a rapid rate. The second resulted from a dose 1 effectiveness against death was less than 65% (that is, more than 32% less effective than dose 2). Recently, effectiveness estimates from the United Kingdom have reported dose 1 effectiveness against death of approximately 80% in adults ≥80 years of age and.^17,18^ In addition, dose 1 effectiveness against hospitalizations, which were a condition for deaths in our model, have been reported at 70-80%, largely in elderly individuals.^17,19^ Examination of additional third wave scenarios showed that, as the severity of our simulated third wave increased, extended intervals could confer benefits at lower effectiveness values as individuals with longer wait times for vaccines faced increasing cumulative risks of infection and onward transmission as well as severe outcomes. Conversely, as the severity of the simulated third wave decreased, extended intervals required higher effectiveness against death after the first dose to reduce deaths as there are fewer deaths to be prevented in a milder resurgence (Supplementary Materials). While the third wave scenarios are not forecasts of how the epidemic will necessarily unfold in Canada, they illustrate how extended intervals provide a strategy to reduce morbidity and mortality when there is an expectation of increasing risks of infection and severe outcomes in the short term. Our model findings can also be extended to other dual dose vaccines such as viral vector vaccines, which have shown early indications of similar performance against severe outcomes.^17,19^ Although our findings reflect population-level effects, it is important to consider the impact of extending the dose interval in subgroups for whom the vaccine may be less effective, such as immunosuppressed individuals.^25,26^

The influence of first-dose effectiveness and duration of protection have similarly been highlighted by other vaccine models that compared extended dosing intervals to no delay for mRNA-vaccination strategies,^27,28^ as well as vaccine models that compared the use of different proportions of the vaccine supply for extended dosing strategies.^29^ Extended intervals up to 24 weeks were preferred given high first dose effectiveness against disease (Moghadas et al: 80%^28^; Jurgens and Lackner: 46.5%^27^), or given limited waning (greater than 18-week duration of protection when first dose effectiveness was low^28^; up to ∼10% waning per month^27^). Using a greater proportion of the supply for extended dosing strategies was preferred even with extreme waning assumptions (e.g., protection drops to zero within 6 weeks of not receiving second dose).^29^ However, we note that these models focused on effectiveness against infection and/or disease as data on effectiveness against other outcomes were limited at the time of those studies.

There are some limitations to the present study. First, our model stratified risk by age and was used to inform broad population-based vaccination strategies but was not designed to examine other high-risk groups such as immunocompromised individuals for whom extended intervals may not be an optimal strategy. Second, we did not consider long-term implications of immune responses, vaccine escape, and variants of concern. However, the third wave simulated in our model and our sensitivity analyses could be considered as a proxy for variant of concern scenarios with higher transmission rates, variable vaccine effectiveness or waning protection. Our sensitivity analyses can inform ongoing evaluation of extended intervals if effectiveness begins to diminish or waning protection accelerates. In addition, we used a simplistic epidemic scenario and did not simulate scenarios of dynamic public health measures that may be deployed to confront a third wave or any subsequent resurgences following implementation of additional public health measures.

Our model adds to the current literature examining different mRNA dose interval strategies with explicit consideration of real-world effectiveness against symptomatic disease, hospitalizations and death. Strategies of extended intervals were examined in the context of the early COVID-19 vaccination campaign in Canada. Further, our model findings can be used to inform ongoing monitoring of extended interval strategies as effectiveness data continue to unfold.

## Conclusion

In conclusion, our modelling is generally consistent with other models, supporting the extension of dose intervals across all age groups for population benefit during a period of constrained supply with a largely un-vaccinated population. Under our base-case scenario and in most sensitivity analyses, extended intervals will reduce symptomatic disease, hospitalizations, and deaths while vaccine supply is constrained.

## Data Availability

Data sources are listed in the manuscript and supplementary material.

## References

1. Public Health Agency of Canada. Statement from the Council of Chief Medical Officers of Health: Implementing COVID-19 Vaccination in Canada — Vaccine Dose Interval [Internet]. 2021 [cited 2021 Mar 11]. Available from: https://www.canada.ca/en/public-health/news/2021/01/statement-from-the-council-of-chief-medical-officers-of-health-implementing-covid-19-vaccination-in-canada--vaccine-dose-interval.html

2. Government of United Kingdom. Optimising the COVID-19 vaccination programme for maximum short-term impact [Internet]. 2020. Available from: https://www.gov.uk/government/publications/prioritising-the-first-covid-19-vaccine-dose-jcvi-statement/optimising-the-covid-19-vaccination-programme-for-maximum-short-term-impact

3. Institut national de, santé publique du Québec. Requested supplement to the notice Strategy for Vaccination Against COVID-19: Postponement of the Second Dose in a Context of Shortage [Internet]. 2020. Available from: https://www.inspq.qc.ca/en/publications/3103

4. Statistics Canada. Table 17-10-0005-01 Population estimates on July 1st, by age and sex.

5. Prem K, Cook AR, Jit M. Projecting social contact matrices in 152 countries using contact surveys and demographic data. PLOS Comput Biol. 2017 Sep 12;13(9):e1005697.

6. Baden LR, El Sahly HM, Essink B, Kotloff K, Frey S, Novak R, et al. Efficacy and Safety of the mRNA-1273 SARS-CoV-2 Vaccine. N Engl J Med. 2020 Dec 30;384:403–16.

7. Polack FP, Thomas SJ, Kitchin N, Absalon J, Gurtman A, Lockhart S, et al. Safety and Efficacy of the BNT162b2 mRNA Covid-19 Vaccine. N Engl J Med. 2020 Dec 10;383(27):2603–15.

8. Voysey M, Costa Clemens SA, Madhi SA, Weckx LY, Folegatti PM, Aley PK, et al. Single-dose administration and the influence of the timing of the booster dose on immunogenicity and efficacy of ChAdOx1 nCoV-19 (AZD1222) vaccine: a pooled analysis of four randomised trials. Lancet. 2021;397(10277):881–91.

9. EKOS Politics. Pandemic, Polarization, and Expectations for Government. [Internet]. 2020. Available from: https://www.ekospolitics.com/index.php/2020/12/pandemic-polarization-and-expectations-for-government/

10. Swan DA, Bracis C, Janes H, Moore M, Matrajt L, Reeves DB, et al. COVID-19 vaccines that reduce symptoms but do not block infection need higher coverage and faster rollout to achieve population impact. medRxiv. 2020 Jan 1;2020.12.13.20248142.

11. Shah AS V, Gribben C, Bishop J, Hanlon P, Caldwell D, Wood R, et al. Effect of vaccination on transmission of COVID-19: an observational study in healthcare workers and their households. medRxiv. 2021 Jan 1;2021.03.11.21253275.

12. Heymann AD, Zacay G, Shasha D, Bareket R, Kadim I, Sikron FH, et al. BNT162b2 Vaccine Effectiveness in Preventing Asymptomatic Infection with SARS-CoV-2 Virus: A Nationwide Historical Cohort Study. Available SSRN https://ssrn.com/abstract=3796868 or https://dx.doi.org/102139/ssrn3796868.

13. Lumley SF, Rodger G, Constantinides B, Sanderson N, Chau KK, Street TL, et al. An observational cohort study on the incidence of SARS-CoV-2 infection and B.1.1.7 variant infection in healthcare workers by antibody and vaccination status. medRxiv. 2021 Jan 1;2021.03.09.21253218.

14. Pawlowski C, Lenehan P, Puranik A, Agarwal V, Venkatakrishnan AJ, Niesen MJM, et al. FDA-authorized COVID-19 vaccines are effective per real-world evidence synthesized across a multi-state health system. medRxiv. 2021 Jan 1;2021.02.15.21251623.

15. Hall V, Foulkes S, Saei A, Andrews N, Oguti B, Charlett A, et al. Effectiveness of BNT162b2 mRNA Vaccine Against Infection and COVID-19 Vaccine Coverage in Healthcare Workers in England, Multicentre Prospective Cohort Study (the SIREN Study). Available SSRN https://ssrn.com/abstract=3790399 or https://dx.doi.org/102139/ssrn3790399.

16. Public Health England. PHE monitoring of the early impact and effectiveness of COVID-19 vaccination in England, March 2021 [Internet]. 2021 [cited 2021 Mar 25]. Available from: https://assets.publishing.service.gov.uk/government/uploads/system/uploads/attachment_data/file/971017/SP_PH__VE_report_20210317_CC_JLB.pdf

17. Bernal JL, Andrews N, Gower C, Stowe J, Robertson C, Tessier E, et al. Early effectiveness of COVID-19 vaccination with BNT162b2 mRNA vaccine and ChAdOx1 adenovirus vector vaccine on symptomatic disease, hospitalisations and mortality in older adults in England. medRxiv. 2021 Jan 1;2021.03.01.21252652.

18. Public Health England. Impact of COVID-19 vaccines on mortality in England. December 2020 to February 2021. [Internet]. 2021 [cited 2003 Jan 20]. Available from: https://assets.publishing.service.gov.uk/government/uploads/system/uploads/attachment_data/file/972592/COVID-19_vaccine_impact_on_mortality_240321.pdf

19. Hyams C, Marlow R, Maseko Z, King J, Ward L, Fox K, et al. Assessing the Effectiveness of BNT162b2 and ChAdOx1nCoV-19 COVID-19 Vaccination in Prevention of Hospitalisations in Elderly and Frail Adults: A Single Centre Test Negative Case-Control Study. Available SSRN https://ssrn.com/abstract=3796835 or https://dx.doi.org/102139/ssrn3796835.

20. Haas EJ, Angulo FJ, McLaughlin JM, Anis E, Singer SR, Khan F, et al. Nationwide Vaccination Campaign with BNT162b2 in Israel Demonstrates High Vaccine Effectiveness and Marked Declines in Incidence of SARS-CoV-2 Infections and COVID-19 Cases, Hospitalizations, and Deaths. Available SSRN https://ssrn.com/abstract=3811387.

21. Dagan N, Barda N, Kepten E, Miron O, Perchik S, Katz MA, et al. BNT162b2 mRNA Covid-19 Vaccine in a Nationwide Mass Vaccination Setting. N Engl J Med. 2021 Feb 24;

22. Tasker JP. Canada to receive 1 million COVID-19 vaccine doses a week starting in April: general [Internet]. CBC News. 2021. Available from: https://www.cbc.ca/news/politics/vaccine-rollout-update-fortin-1.5872766

23. Tasker JP. More Pfizer shots will arrive in 2nd quarter than originally planned: Trudeau [Internet]. CBC News. 2021. Available from: https://www.cbc.ca/news/politics/more-pfizer-shots-trudeau-1.5912209

24. Little N. COVID-19 Tracker Canada [Internet]. 2020 [cited 2021 Mar 23]. Available from: https://covid19tracker.ca

25. Boyarsky BJ, Werbel WA, Avery RK, Tobian AAR, Massie AB, Segev DL, et al. Immunogenicity of a Single Dose of SARS-CoV-2 Messenger RNA Vaccine in Solid Organ Transplant Recipients. JAMA. 2021 Mar 15;

26. Monin-Aldama L, Laing AG, Muñoz-Ruiz M, McKenzie DR, del Molino del Barrio I, Alaguthurai T, et al. Interim results of the safety and immune-efficacy of 1 versus 2 doses of COVID-19 vaccine BNT162b2 for cancer patients in the context of the UK vaccine priority guidelines. medRxiv. 2021 Jan 1;2021.03.17.21253131.

27. Jurgens GT, Lackner K. Modelled Optimization of SARS-Cov-2 Vaccine Distribution: an Evaluation of Second Dose Deferral Spacing of 6, 12, and 24 weeks. medRxiv. 2021 Jan 1;2021.02.28.21252638.

28. Moghadas SM, Vilches TN, Zhang K, Nourbakhsh S, Sah P, Fitzpatrick MC, et al. Evaluation of COVID-19 vaccination strategies with a delayed second dose. medRxiv. 2021 Jan 1;2021.01.27.21250619.

29. Tuite AR, Zhu L, Fisman DN, Salomon JA. Alternative Dose Allocation Strategies to Increase Benefits From Constrained COVID-19 Vaccine Supply. Ann Intern Med. 2021 Jan 5;

30. Zhao S. Estimating the time interval between transmission generations when negative values occur in the serial interval data: using COVID-19 as an example. Math Biosci Eng. 2020;17(4):3512–9.

31. He X, Lau EHY, Wu P, Deng X, Wang J, Hao X, et al. Temporal dynamics in viral shedding and transmissibility of COVID-19. Nat Med. 2020;26(5):672–5.

32. Public Health Agency of Canada. Canada COVID-19 Weekly Epidemiology Report 17 January to 23 January 2021 [Internet]. 2021. Available from: https://www.canada.ca/content/dam/phac-aspc/documents/services/diseases/2019-novel-coronavirus-infection/surv-covid19-weekly-epi-update-20210129-eng.pdf

33. Canadian Institute for Health Information. COVID-19 Hospitalization and Emergency Department Statistics, 2019–2020 and 2020–2021. Ottawa, ON: CIHI; 2020.

