## Supplementary Materials for "Modelling the impact of extending dose intervals for COVID-19 vaccines in Canada"

### Model equations

#### *Force of infection*

Each compartment is subdivided into separate components for each age group (16 age groups in five-year intervals), represented by the subscript *i*. Note: a table of parameter definitions is available at the end of this supplement.

The force of infection ($\lambda_i$) among susceptible individuals (in age group *i*) is conceptualized as a function of contacts with asymptomatic/pre-symptomatic and mild symptomatic individuals ($\lambda_i'$) and with severe symptomatic individuals ($\lambda_i''$), who are assumed to self-isolate. Infection rate is parameterized with a contact rate between individuals in age group *i* and *j* ($cr_{ij}$), probability of transmission given contact ($\beta$), and a time-dependent reduction associated with public health measures and physical distancing ($f(t)$). A separate contact rate with individuals who have severe symptoms ($cro_{ij}$) was used to represent the assumption that all severe cases self-isolate and is assumed to be 25% of projected home contacts.

$$\lambda_i = \lambda_i' + \lambda_i''$$

$$\lambda_i' = \sum_{j=1}^{16} \frac{1}{N_j} \beta cr_{ij} \cdot rrpd_t \left(A_j + Av_j + P_j + Pv_j + Pv_j' + Pv_j'' + M_j + Mv_j\right)$$

$$\lambda_i'' = \sum_{j=1}^{16} \frac{1}{N_j} \beta cro_{ij} \left(X_j + Xv_j + Xv_j' + Xv_j''\right)$$

*Natural history of infection and disease*

$$\frac{dS_i}{dt} = -\lambda_i S_i - \frac{S_i}{S_i + R_i}\Omega_{dose1} + \xi'' Vx''_i + w(R_i + Rv_i) - \chi(S_i - S_{i-1})$$

$$\frac{dE_i}{dt} = \lambda_i S_i - \rho E_i - \chi(E_i - E_{i-1})$$

$$\frac{dA_i}{dt} = \alpha\rho E_i - \left({\delta_p}^{-1} + \gamma^{-1}\right)^{-1} A_i - \chi(A_i - A_{i-1})$$

$$\frac{dP_i}{dt} = (1-\alpha)\rho E_i - \delta_p P_i - \chi(P_i - P_{i-1})$$

$$\frac{dM_i}{dt} = (1-\kappa)\delta_p P_i - \gamma M_i - \chi(M_i - M_{i-1})$$

$$\frac{dX_i}{dt} = \kappa\delta_p P_i - \delta_x X_i - \chi(X_i - X_{i-1})$$

$$\frac{dH_i}{dt} = \delta_x X_i - \frac{(1-\mu_h)}{losr_h} H_i - \frac{\mu_h}{losd_h} H_i - \chi(H_i - H_{i-1})$$

$$\frac{dR_i}{dt} = \left({\delta_p}^{-1} + \gamma^{-1}\right)^{-1} A_i + \gamma M_i + \frac{(1-\mu_h)}{losr_h} H_i - \frac{R_i}{S_i + R_i}\Omega_{dose1} - wR_i - \chi(R_i - R_{i-1})$$

$$\frac{dD_i}{dt} = \frac{\mu_h}{losd_h}(H_i + Hv_i) + (1 - ve'_{death})\frac{\mu_h}{losd_h} Hv'_i + (1 - ve''_{death})\frac{\mu_h}{losd_h} Hv''_i$$

*Vaccination*

We conceptualize the maximum daily vaccine uptake as a function of the current vaccine supply, $Z$, and the vaccination capacity, $vaccrate_{max}$, and the number of individuals eligible for vaccination. The maximum daily number of doses available for administration, $\Omega$, is the lesser of current vaccine supply, $Z$, and the vaccination capacity, $vaccrate_{max}$. We also proportioned the daily vaccine supply over the week. The maximum daily doses is prioritized for all individuals who are waiting for dose 2 and any remaining doses are then allocated to individuals eligible for dose 1.

$$\frac{dZ}{dt} = -\Omega_{dose1} - \Omega_{dose2}$$

$$\Omega = min\left(\frac{Z}{days\ remaining\ in\ supply\ week}, vaccrate_{max}\right)$$

$$\Omega_{dose1} = min\left(S + R, max\left(0, \Omega - Vq'_i - Vs'_i - Vrq'_i\right)\right)$$

$$\Omega_{dose2} = min(\Omega, Vq'_i + Vs'_i + Vrq'_i)$$

Vaccinated individuals are initially subject to the same force of infection as unvaccinated individuals. If infected, individuals can move to a latent, vaccinated compartment ($Ev_i$). Protection from symptomatic COVID-19 is applied against the fraction of individuals who transition to the pre-symptomatic compartment ($1 - ve'_{symp}$, $1 - ve''_{symp}$). Uninfected individuals transition to a partially protected compartment where they wait for the second dose for the remaining time of the base case interval ($tt_{dose2}$). In scenarios where extended intervals are considered, individuals wait in a queued compartment ($Vq'_i$) before receiving their second dose. We assumed that waning protection does not begin until after the base case interval has elapsed ($tt_{dose2}$). Individuals whose protection wanes, move to a separate compartment ($Vs'_i$) where they are subject to the same force of infection as unvaccinated individuals. Individuals with waned protection receive a second dose if they remain uninfected. The effect of the second dose begins after a pre-set period prior to which vaccinated individuals are subject to a reduced force of infection conferred by first dose protection.

*First dose*

$$\frac{dVn_i}{dt} = \frac{S_i}{S_i + R_i}\Omega_{dose1} - \psi' Vn_i - \lambda_i Vn_i - \chi(Vn_i - Vn_{i-1})$$

$$\frac{dV'_i}{dt} = \psi' Vn_i - \left(1 - ve'_{inf}\right)\lambda_i V'_i - \frac{V'_i}{({tt_{dose2}}^{-1} - \psi'^{-1})} - \chi(V'_i - V'_{i-1})$$

$$\frac{dVq'_i}{dt} = \frac{V'_i}{{tt_{dose2}}^{-1} - \psi'^{-1}} - \frac{Vq'_i}{Vq'_i + Vs'_i + Vrq'_i}\tau_1\Omega_{dose2} - \left(1 - ve'_{inf}\right)\lambda_i Vq'_i - \xi' Vq'_i - \chi(Vq'_i - Vq'_{i-1})$$

$$\frac{dVs'_i}{dt} = \xi' Vq'_i - \frac{Vs'_i}{Vq'_i + Vs'_i + Vrq'_i}\tau_2\Omega_{dose} - \lambda_i Vs'_i - \chi(Vs'_i - Vs'_{i-1})$$

$$\frac{dVr'_i}{dt} = \frac{R_i}{S_i + R_i}\Omega_{dose1} - \frac{Vr'_i}{tt_{dose2}} - \chi(Vr'_i - Vr'_{i-1})$$

$$\frac{dVrq'_i}{dt} = \frac{Vr'_i}{tt_{dose2}} - \frac{Vrq'_i}{Vq'_i + Vs'_i + Vrq'_i}\tau_1\Omega_{dose2} - \chi(Vrq'_i - Vrq'_{i-1})$$

*Second dose*

$$\frac{dV''_i}{dt} = \frac{Vq'_i}{Vq'_i + Vs'_i + Vrq'_i}\tau_1\Omega_{dose2} - \psi''V''_i - \left(1 - ve'_{inf}\right)V''_i - \chi(V''_i - V''_{i-1})$$

$$\frac{dVs''_i}{dt} = \frac{Vs'_i}{Vq'_i + Vs'_i + Vrq'_i}\tau_2\Omega_{dose2} - \psi''Vs''_i - \lambda_i Vs''_i - \chi(Vs''_i - Vs''_{i-1})$$

$$\frac{dVx''_i}{dt} = \psi''(V''_i + Vs''_i) - \xi''Vx''_i - \left(1 - ve''_{inf}\right)\lambda_i Vx''_i - \chi(Vx''_i - Vx''_{i-1})$$

$$\frac{dVr''_i}{dt} = \frac{Vrq'_i}{Vq'_i + Vs'_i + Vrq'_i}\tau_1\Omega_{dose2} - \psi''Vr''_i - \chi(Vr''_i - Vr''_{i-1})$$

*Vaccinated, Infected*

$$\frac{dEv_i}{dt} = \lambda_i(Vn_i + Vs'_i + Vs''_i) - \rho Ev_i - \chi(Ev_i - Ev_{i-1})$$

$$\frac{dEv'_i}{dt} = \left(1 - ve'_{inf}\right)\lambda_i(V'_i + Vq'_i + V''_i) - \rho Ev'_i - \chi(Ev'_i - Ev'_{i-1})$$

$$\frac{dEv''_i}{dt} = \left(1 - ve''_{inf}\right)\lambda_i Vx''_i - \rho Ev''_i - \chi(Ev''_i - Ev''_{i-1})$$

We define an adjustment factor, $xf_{symp}$, to ensure that the total transition rate out of the latent compartment does not change with vaccination.

$$\left(1 - ve_{symp}\right)(1 - \alpha) + xf_{symp}\alpha\rho = \rho$$

$$1 - \alpha - ve_{symp} + \alpha ve_{symp} + xf_{symp}\alpha = 1$$

$$xf_{symp} = \frac{\alpha + ve_{symp} - \alpha ve_{symp}}{\alpha}$$

$$\frac{dAv_i}{dt} = \alpha\rho\left[Ev_i + \frac{ve'_{symp} + \alpha - ve'_{symp}\alpha}{\alpha}Ev'_i + \frac{ve''_{symp} + \alpha - ve''_{symp}\alpha}{\alpha}Ev''_i\right] - \left(\delta_p{}^{-1} + \gamma^{-1}\right)^{-1}Av_i - \chi(Av_i - Av_{i-1})$$

$$\frac{dPv_i}{dt} = (1 - \alpha)\rho Ev_i - \delta_p Pv_i - \chi(Pv_i - Pv_{i-1})$$

$$\frac{dPv_i'}{dt} = \left(1 - ve_{symp}'\right)(1-\alpha)\rho Ev_i' - \delta_p Pv_i' - \chi(Pv_i' - Pv_{i-1}')$$

$$\frac{dPv_i''}{dt} = \left(1 - ve_{symp}''\right)(1-\alpha)\rho Ev_i'' - \delta_p Pv_i'' - \chi(Pv_i'' - Pv_{i-1}'')$$

We define an adjustment factor, $xf_{hosp}$, to ensure that the total transition rate out of the pre-symptomatic compartment and into the mild/moderate and severe compartments does not change with vaccination.

$$xf_{hosp}(1-\kappa)\delta_p + \left(1 - ve_{hosp}\right)\kappa\delta_p = \delta_p$$

$$xf_{hosp} - xf_{hosp}\kappa + \kappa - ve_{hosp}\kappa = 1$$

$$xf_{hosp} = \frac{1 - \kappa + ve_{hosp}\kappa}{1-\kappa}$$

$$\frac{dMv_i}{dt} = (1-\kappa)\delta_p\left(Pv_i + \frac{1-\kappa+ve_{hosp}'\kappa}{1-\kappa}Pv_i' + \frac{1-\kappa+ve_{hosp}''\kappa}{1-\kappa}Pv_i''\right) - \gamma Mv_i - \chi(Mv_i - Mv_{i-1})$$

$$\frac{dXv_i}{dt} = \kappa\delta_p Pv_i - \delta_x Xv_i - \chi(Xv_i - Xv_{i-1})$$

$$\frac{dXv_i'}{dt} = \left(1 - ve_{hosp}'\right)\kappa\delta_p Pv_i' - \delta_x Xv_i' - \chi(Xv_i' - Xv_{i-1}')$$

$$\frac{dXv_i''}{dt} = \left(1 - ve_{hosp}''\right)\kappa\delta_p Pv_i'' - \delta_x Xv_i'' - \chi(Xv_i'' - Xv_{i-1}'')$$

$$\frac{dHv_i}{dt} = \delta_x Xv_i - \frac{(1-\mu_h)}{losr_h}Hv_i - \frac{\mu_h}{losd_h}Hv_i - \chi(Hv_i - Hv_{i-1})$$

We define an adjustment factor, $xf_{death}$, to ensure that the total transition rate out of the hospitalized compartment (survivors + non-survivors) does not change with vaccination.

$$xf_{death}\frac{1-\mu_h}{losr_h} + (1 - ve_{death})\frac{\mu_h}{losd_h} = \frac{1-\mu_h}{losr_h} + \frac{\mu_h}{losd_h}$$

$$xf_{death}(1-\mu_h)losd_h + (1 - ve_{death})\mu_h losr_h = (1-\mu_h)losd_h + \mu_h losr_h$$

$$xf_{death} = \frac{(1-\mu_h)losd_h + \mu_h losr_h - \mu_h losr_h + ve_{death}\mu_h losr_h}{(1-\mu_h)losd_h}$$

$$xf_{death} = 1 + \frac{ve_{death}\mu_h losr_h}{(1-\mu_h)losd_h}$$

$$\frac{dHv_i'}{dt} = \delta_x Xv_i' - \left(1 + \frac{ve_{death}'\mu_h losr_h}{(1-\mu_h)losd_h}\right)\left(\frac{1-\mu_h}{losr_h}\right)Hv_i' - (1 - ve_{death}')\frac{\mu_h}{losd_h}Hv_i' - \chi(Hv_i' - Hv_i'')$$

$$\frac{dHv_i''}{dt} = \delta_x Xv_i'' - \left(1 + \frac{ve_{death}''\mu_h losr_h}{(1-\mu_h)losd_h}\right)\left(\frac{1-\mu_h}{losr_h}\right)Hv_i'' - (1 - ve_{death}'')\frac{\mu_h}{losd_h}Hv_i'' - \chi(Hv_i'' - Hv_{i-1}'')$$

$$\frac{dRv_i}{dt} = \left(\delta_p{}^{-1} + \gamma^{-1}\right)^{-1} Av_i + \gamma Mv_i + \frac{(1-\mu_h)}{losr_h}Hv_i + \left(1 + \frac{ve_{death}'\mu_h losr_h}{(1-\mu_h)losd_h}\right)\left(\frac{1-\mu_h}{losr_h}\right)Hv_i' + \left(1 + \frac{ve_{death}''\mu_h losr_h}{(1-\mu_h)losd_h}\right)\left(\frac{1-\mu_h}{losr_h}\right)Hv_i'' + \psi'' Vr''_i - wRv_i - \chi(Rv_i - Rv_{i-1})$$

*Table S1. Compartment definitions*

| Compartment | Definition |
|---|---|
| $S$ | Susceptible |
| $E$ | Infected (latent) |
| $A$ | Infectious, asymptomatic |
| $P$ | Pre-symptomatic |
| $M$ | Symptomatic (mild) |
| $X$ | Symptomatic (severe) |
| $H$ | Hospitalized |
| $R$ | Recovered |
| $D$ | Dead |
| $Vn$ | Vaccinated, first dose, no effect |
| $V'$ | Vaccinated, first dose, first dose effect |
| $Vq'$ | Vaccinated, first dose, first dose effect, queued for second dose |
| $Vs'$ | Vaccinated, first dose, first dose waned protection, queued for second dose |
| $V''$ | Vaccinated, second dose, first dose efect |
| $Vs''$ | Vaccinated, second dose, first dose waned protection |
| $Vx''$ | Vaccinated, second dose, second dose effect |
| $Vr'$ | Vaccinated, first dose, recovered from previous infection |
| $Vrq'$ | Vaccinated, queued for second dose, recovered from previous infection |
| $Vr''$ | Vaccinated, second dose, recovered from previous infection |
| $Ev$ | Vaccinated, no protection, infected (latent) |
| $Ev'$ | Vaccinated, first dose protection, infected (latent) |
| $Ev''$ | Vaccinated, second dose protection, infected (latent) |
| $Av$ | Vaccinated, infectious, asymptomatic |
| $Pv$ | Vaccinated, no protection, pre-symptomatic |
| $Pv'$ | Vaccinated, first dose protection, pre-symptomatic |
| $Pv''$ | Vaccinated, second dose protection, pre-symptomatic |
| $Mv$ | Vaccinated, symptomatic (mild) |
| $Xv$ | Vaccinated, no protection, symptomatic (severe) |
| $Xv'$ | Vaccinated, first dose protection, symptomatic (severe) |
| $Xv''$ | Vaccinated, second dose protection, symptomatic (severe) |
| $Hv$ | Vaccinated, no protection, hospitalized |
| $Hv'$ | Vaccinated, first dose protection, hospitalized |
| $Hv''$ | Vaccinated, second dose protection, hospitalized |
| $Rv$ | Vaccinated, recovered |

### Model calibration

We calibrated model parameters to reported COVID-19 hospitalizations and deaths in the province of Ontario from March 16 up to December 18, 2020. We used four calibration targets: daily hospitalizations, daily deaths, cumulative hospitalizations by age group, and cumulative deaths by age group. Aggregated daily hospitalization data were obtained from Ontario's case and contact management (CCM Plus) database.[1] Aggregated daily deaths were obtained from the Ontario Data Catalogue.[2] We used age-stratified proportions of hospitalizations and deaths in Canada, reported by the Public Health Agency of Canada, to infer cumulative hospitalizations by age. To derive cumulative deaths by age, we used the proportion of deaths in Canada for ages 0-64 years and used the proportion of deaths by age for individuals 65 years and older reported by Public Health Ontario, stratified by long-term care status.[3] We applied the proportion of deaths by age group against the total deaths on December 18, 2020 to derive the cumulative deaths by age. Table S2 lists the distributions of hospitalizations and deaths by age that were used as calibration targets.

*Table S2. Distributions of hospitalizations and deaths by age inferred as calibration targets*

| Age | Hospitalizations (%) | Deaths (%) |
|---|---|---|
| 0-19 | 1.5 | 0 |
| 20-29 | 3.1 | 0.3 |
| 30-39 | 5.1 | 0.8 |
| 40-49 | 7.0 | 1.8 |
| 50-59 | 12.0 | 6.4 |
| 60-69 | 16.8 | 10.7 |
| 70-74 | 10.5 | 12.2 |
| 75+ | 44.0 | 67.8 |

To infer parameters of the transmission model, we fitted a time-dependent parameter ($rrpd_t$) that modulated the force of infection to represent the collective effect of public health measures and compliance with physical and social distancing. We fitted values for $rrpd_t$ for four different periods from March 16 to December 18, 2020 (Table S3). For the periods covering March 16 to July 17, 2020 (lockdown and limited recovery) and July 17 to September 8, 2020 (Stage 3 recovery), we allowed $rrpd_t$ to change linearly with time until it reached the first and second fitted values. We also calibrated the daily rates of decrease, $tf_1$, and increase, $tf_2$, in $rrpd_t$. For the remaining periods, $rrpd_t$ was subject to a one-time change at the beginning of each period. We describe this calibration of $rrpd_t$ with the following function:

$$f(t) = \begin{cases} 1, t < 75 \\ max(1 - (t - 75)tf_1, rrpd1), 75 \leq t < 198 \\ min(rrpd_1 + (t - 198)tf_2, rrpd2, 198 \leq t < 251) \\ rrpd3, 251 \leq t < 283 \\ rrpd4, 283 \leq t < 353 \end{cases}$$

In addition, we searched for a time point to initialize the model with 100 infections in the 40-year old latent compartment.

*Table S3. Fitting time-dependent values for $rrpd_t$*

| **Period** | **Rationale** | **$rrpd_t$** |
|---|---|---|
| March 16 – July 17 | Lockdown, Stage 1 recovery, Stage 2 recovery | $rrpd_t$ decreased linearly with time to the first fitted value |
| July 17 – September 8 | Stage 3 recovery | $rrpd_t$ increased linearly with time to the second fitted value |
| September 8 – October 10 | Partial re-opening of schools | A one-time increase in $rrpd_t$ (third fitted value) |
| October 10 – December 18 | Modified Stage 2 | A one-time decrease in $rrpd_t$ (fourth fitted value) |

We used a Differential-Evolution Monte Carlo Markov Chain (DE-MCMC) algorithm,[4,5] using 12 Markov chains to sample the target distribution. The algorithm was run with a burn-in period until the potential scale reduction factor fell below 1.05 for all parameters. 10,000 samples were then drawn for each chain following the burn-in period. We note that we used a smaller number of chains than recommended in the literature, but chose 12 chains as a trade-off in computational cost. Thus, it is possible that the posterior distributions may not have been fully sampled due to suboptimal mixing between chains.

We constrained the parameter space based on published and pre-published COVID-19 literature and other public reports relevant to the Canadian epidemic (Table S4). The model was fitted to daily hospitalizations and deaths using a Poisson distribution and to cumulative hospitalizations and deaths using a binomial distribution. All model parameters (fitted and fixed) are listed in Table S5. We used informative priors for the asymptomatic proportion, latent period, and pre-symptomatic period to reflect a stronger assumption about these values and to anchor the posterior distributions for remaining parameters around these assumptions. We used uninformative priors for the transmission coefficient, mild/moderate symptom onset to recovery, severe symptom onset to hospitalization, hospital length of stay (non-survivors), age-dependent probabilities of severe/hospitalization, age-dependent probabilities of death, relative reduction in transmission due to public health measures and physical distancing, and rate of change in $rrpd_t$.

*Table S4. Prior and posterior distributions for fitted parameters*

| **Parameter** | **Parameter space** | **Prior** | **Posterior** | **Rationale** |
|---|---|---|---|---|
| Transmission coefficient ($\beta$) | 0 – 1 | Beta (1,1) | Beta (12824, 557000) | N/A |
| Proportion asymptomatic ($\alpha$) | 0.1 – 0.25 | Beta (8, 32) | Beta (1452, 8054) | Byambasuren et al.[6] |
| Latent period ($\frac{1}{\rho}$) | 3 – 5 | Lognormal (1.194, 0.234) | Lognormal (1.376, 0.012) | Zhao et al.[7] |
| Pre-symptomatic period ($\frac{1}{\delta_p}$) | 1 – 3 | Lognormal (0.663, 0.246) | Lognormal (0.907, 0.029) | He et al.[8] |
| Mild/moderate symptom onset to recovery ($\frac{1}{\gamma}$) | 2 – 8 | Uniform (2, 8) | Lognormal (1.631, 0.009) | He et al.[8] |
| Severe symptom onset to hospitalization ($\frac{1}{\delta_x}$) | 3 – 10 | Uniform (3, 10) | Lognormal (1.740, 0.012) | PHAC Weekly Epidemiological Report 17 January to 23 January 2021[9] |
| Length of hospital stay, non-survivors ($losd_h$) | 10 – 30 | Uniform (10, 30) | Lognormal (2.988, 0.003) | CIHI[10] |
| Probability of severe/hospitalization \| infected ($\kappa$) | 0 – 1 | Beta (1,1) | 0-19: Beta (171, 132658)<br>20-29: Beta (273, 65590)<br>30-39: Beta (447, 56249)<br>40-49: Beta (681, 62457)<br>50-59: Beta (1242, 62888)<br>60-69: Beta (1863, 29100)<br>70-74: Beta (1259, 10257)<br>75+: Beta (1217, 1081) | Starting values were chosen from a range based on PHAC surveillance data |
| Probability of death \| hospitalized ($\mu_h$) | 0 – 1 | Beta (1, 1) | 0-19: Beta (35, 5397)<br>20-29: Beta (3, 159)<br>30-39: Beta (131, 2002)<br>40-49: Beta (30, 362) | Starting values were chosen from a range based on PHAC |

| | | | | |
|---|---|---|---|---|
| | | | 50-59: Beta (364, 1570)<br>60-69: Beta (210, 714)<br>70-74: Beta (6529, 9573)<br>75+: Beta (8705, 7818) | surveillance data |
| Relative reduction in transmission ($rrpd_t$) | 0 – 1 | Beta (1, 1) | 03/16 – 07/17: Beta (9717, 22051)<br>07/17 – 09/08: Beta (4518, 6581)<br>09/08 – 10/10: Beta (2947, 1948)<br>10/10 – 12/18: Beta (5654, 6424) | |
| Daily rate of decrease in $rrpd_t$ ($tf_1$) | 0 – 1 | Uniform (0, 1) | 03/16 – 07/17: Lognormal (-2.675, 0.008) | Starting values were chosen from a range based on an assumption that the rate would not exceed 10% per day |
| Daily rate of increase in $rrpd_t$ ($tf_2$) | 0 – 1 | Uniform (0, 1) | 07/17 – 09/11: Lognormal (-4.456, 0.072) | Starting values were chosen from a range based on an assumption that the rate would not exceed 2% per day |
| Time of model initialization (days since January 1, 2020) | 20 – 31 | Uniform (20, 31) | Lognormal (3.283, 0.012) | |

Figures S1-S2 show the results of the fitted model against observed daily hospital admissions and deaths in Ontario (up to December 18, 2020). The model fit well to hospital admissions. The model showed good fit to deaths in the latter half of the year but did not fit as well in the first wave. We believe this may be related to the assumptions used to infer the cumulative deaths by age in the absence of long-term care deaths by age group. The distribution of deaths in older age groups may be different in the latter half of the year than in the first wave and may be contributing to a less optimal fit in the first wave. In addition, dates of long-term care deaths may not have been precisely reported in the first wave, which may affect our estimated deaths in the community (derived by subtracting long-term care deaths from total deaths). However, we believe it is more important for the model to provide a reasonable representation of hospitalizations and deaths over a more recent period. Figures S3-S4 show the results of the fitted model to the target cumulative hospitalizations and deaths by age group up to December 18, 2020. The fitted model produced a distribution of hospitalizations and deaths across ages closely to our calibration targets. We also note a study that estimated susceptibility to infection by age in Ontario, Canada that, at least qualitatively, indicates our fitted estimates of susceptibility to severe disease are similar when considering our asymptomatic fraction.[11]

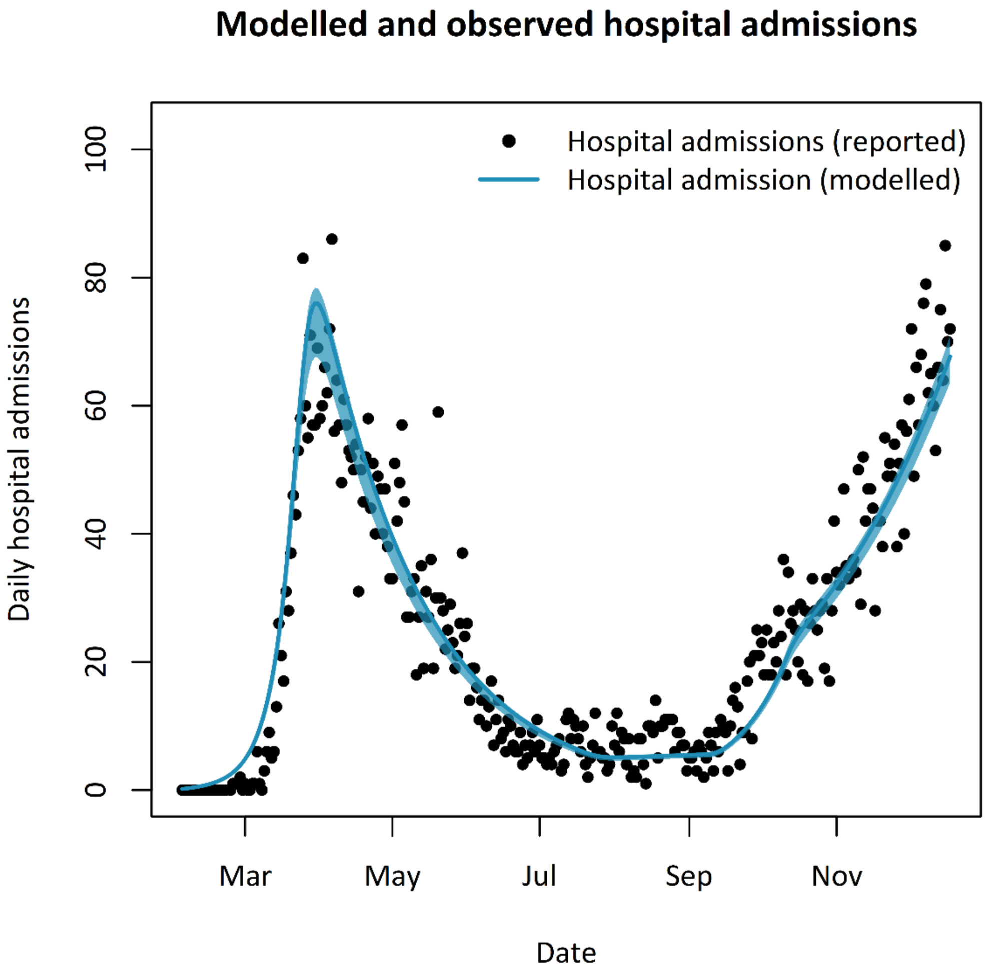


*Figure S1. Fitted hospitalizations: model vs observation*

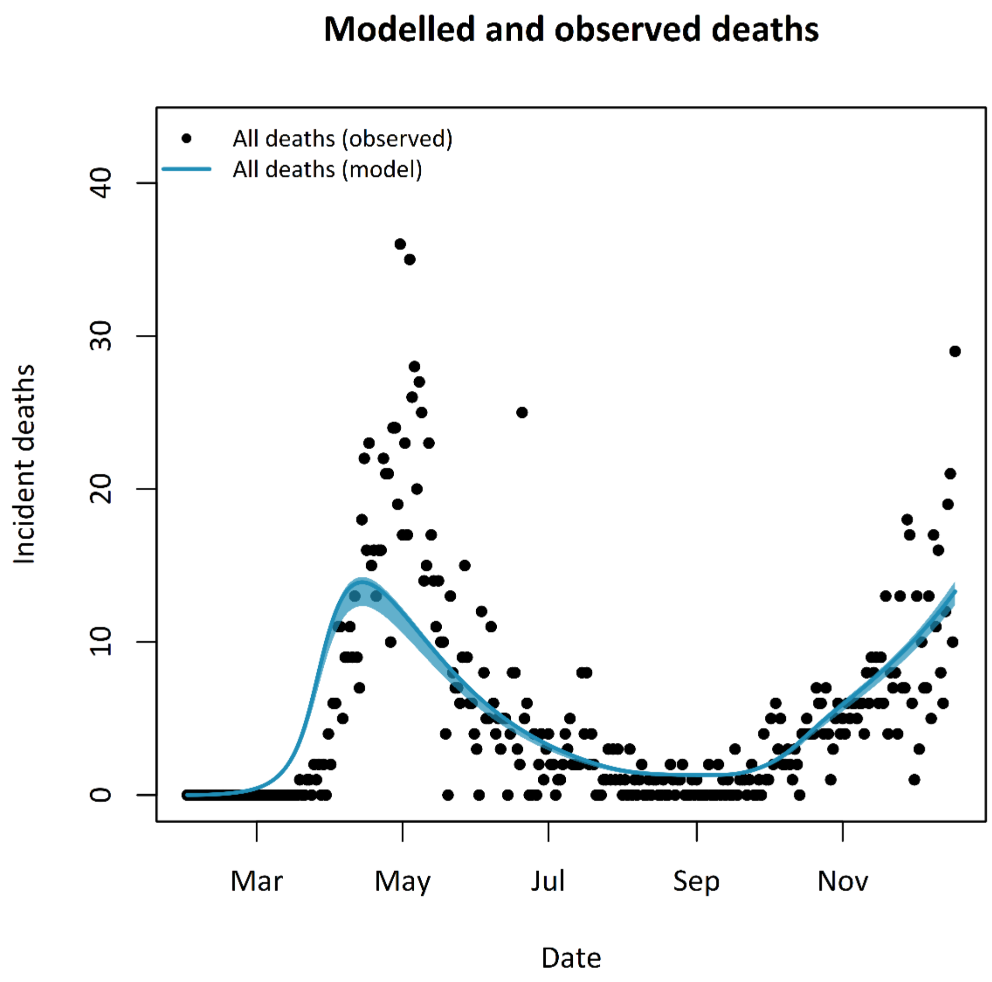


*Figure S2. Fitted deaths: model vs observation.*

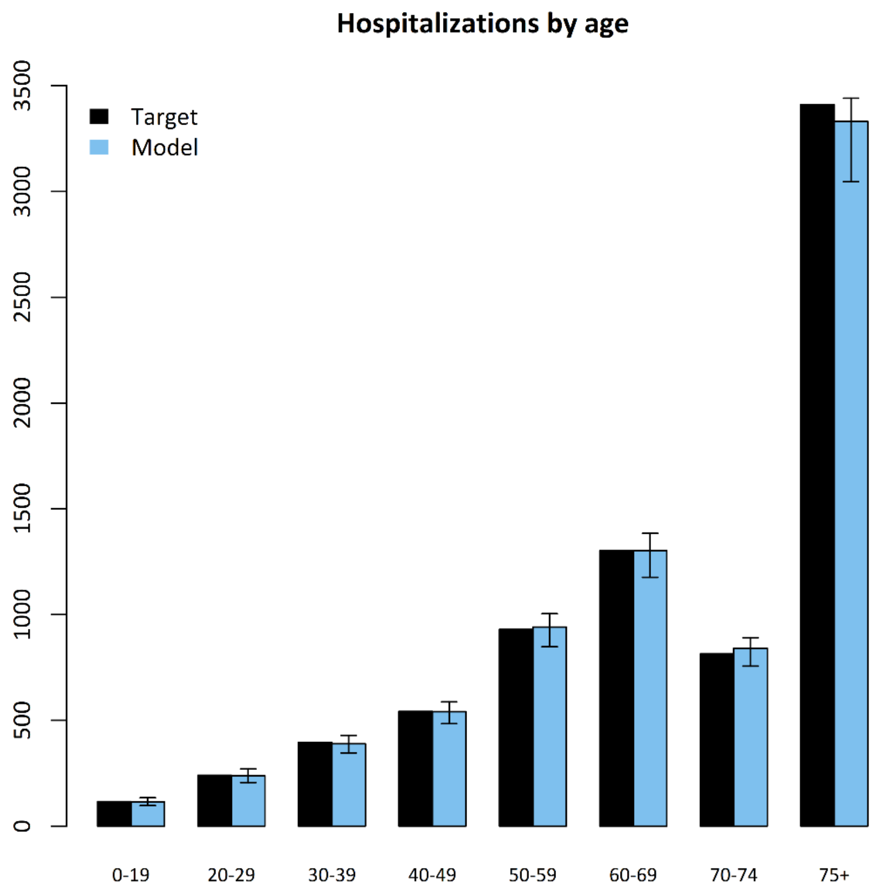


*Figure S3. Cumulative hospitalizations by age up to December 18, 2020.*

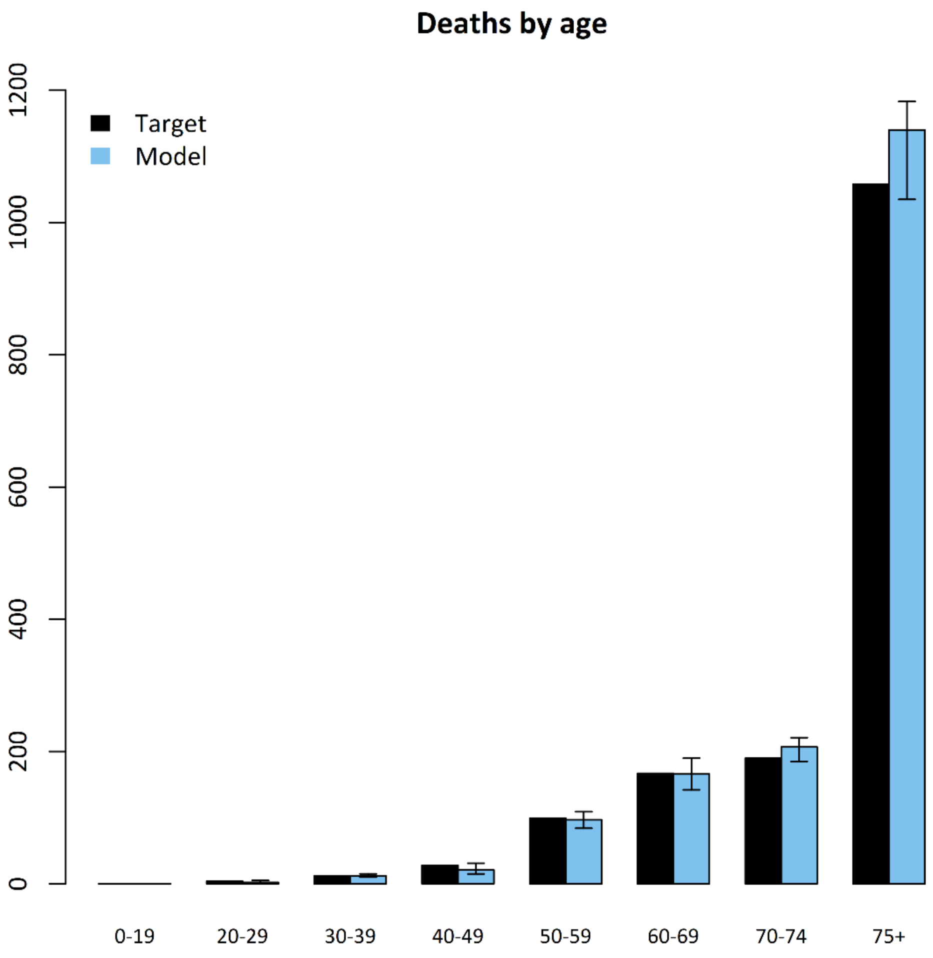


*Figure S4. Cumulative deaths by age (excluding long-term care).*

*Table S5. Model parameters*

| Parameter | Definition | Value | Source |
|---|---|---|---|
| $\beta$ | Probability transmission \| contact | 0.0225 | Fitted |
| $cr_{ij}$ | Contact matrix | See reference | Prem et al.[12] |
| $cro_{ij}$ | Contact matrix \| isolated | Used home contacts from Prem et al. Assumed 25% of home contacts. | Prem et al.[12] |
| $rrpd_t$ | Relative transmission rate associated with public health measures and physical distancing | 03/16 – 07/17: 0.306<br>07/17 – 09/11: 0.407<br>09/11 – 10/10: 0.602<br>10/10 – 12/18: 0.468 | Fitted |
| $tf_1$ | Linear decrease in $rrpd_t$ during period of 03/16 – 07/17 | 0.0689 | Fitted |
| $tf_2$ | Linear increase in $rrpd_t$ during period of 07/17 – 09/11 | 0.0116 | Fitted |
| $\alpha$ | Proportion of asymptomatic infections | 0.152 | Fitted |
| $\frac{1}{\rho}$ | Latent period (days) | 3.96 | Fitted |
| $\frac{1}{\delta_p}$ | Pre-symptomatic period (days) | 2.48 | Fitted |
| $\frac{1}{\gamma}$ | Mild/moderate symptom onset to recovery (days) | 5.11 | Fitted |
| $\frac{1}{\delta_x}$ | Severe symptom onset to hospitalization (days) | 5.70 | Fitted |
| $\kappa$ | Proportion of severe infection \| symptomatic infection | 0-19: 0.0013<br>20-29: 0.0042<br>30-39: 0.0079<br>40-49: 0.0108<br>50-59: 0.0194<br>60-69: 0.0602<br>70-74: 0.1093<br>75+: 0.5298 | Fitted |
| $losr_h$ | Hospital length of stay (survivors) (days) | 11 | PHAC Weekly Report[9] |
| $losd_h$ | Hospital length of stay (non-survivors) (days) | 19.84 | Fitted |
| $\mu_h$ | Probability of death \| hospitalized | 0-19: 0.0064 | Fitted |

| | | | |
|---|---|---|---|
| | | 20-29: 0.0180<br>30-39: 0.0614<br>40-49: 0.0767<br>50-59: 0.1880<br>60-69: 0.2273<br>70-74: 0.4055<br>75+: 0.5268 | |
| $ve'_{inf}$ | First dose effectiveness vs infection | 90% of $ve'_{symp}$<br>50% of $ve'_{symp}$<br>(conservative scenario) | Informed by Heymann et al,[13] Shah et al,[14] Amit et al,[15] Pawlowski et al[16] |
| $ve''_{inf}$ | Second dose effectiveness vs infection | 90% of $ve'_{symp}$<br>50% of $ve'_{symp}$<br>(conservative scenario) | Informed by Heymann et al.,[13] Shah et al.,[14] Amit et al.,[15] Pawlowski et al.,[16] Dagan et al.,[17] Haas et al.[18] |
| $ve'_{symp}$ | First dose effectiveness vs symptomatic disease given infection | $1-\frac{1-VE_{disease,dose\ 1}}{1-ve'_{inf}}$ | Overall effectiveness values in main text |
| $ve''_{symp}$ | First dose effectiveness vs symptomatic disease after second dose | $1-\frac{1-VE_{disease,dose\ 2}}{1-ve''_{inf}}$ | Overall effectiveness values in main text |
| $ve'_{hosp}$ | First dose effectiveness vs hospitalization given symptomatic disease | $1-\frac{1-VE_{hosp,dose\ 1}}{1-VE_{disease,dose1}}$ | Overall effectiveness values in main text |
| $ve''_{hosp}$ | Second dose effectiveness vs hospitalization \| symptomatic disease | $1-\frac{1-VE_{hosp,dose\ 2}}{1-VE_{disease,dose2}}$ | Overall effectiveness values in main text |
| $ve'_{death}$ | First dose effectiveness vs death \| hospitalization | $1-\frac{1-VE_{death,dose\ 1}}{1-VE_{hosp,dose1}}$ | Overall effectiveness values in main text |
| $ve''_{death}$ | Second dose effectiveness vs death \| hospitalization | $1-\frac{1-VE_{death,dose\ 2}}{1-VE_{hosp,dose2}}$ | Overall effectiveness values in main text |
| $\frac{1}{\psi'}$ | Time to efficacy after first dose (days) | 14 | Baden et al.[19] Polack et al.[20] |

| | | | |
|---|---|---|---|
| $\frac{1}{\psi''}$ | Time to efficacy after second dose (days) | 7 | Baden et al.[19]<br>Polack et al.[20] |
| $\frac{1}{\tau_1}$ | Extended dosing interval (days). If no extension to dosing interval, $\tau_1$=1. | | Varied by scenario |
| $\frac{1}{\tau_2}$ | max(1, duration of protection – extended dosing interval) | | |
| $\frac{1}{\zeta'}$ | Duration of vaccine protection after first dose (years) | 2 | Assumption |
| $\frac{1}{\zeta''}$ | Duration of vaccine protection after second dose | Not implemented in model | |
| $\frac{1}{w}$ | Duration of acquired immunity | Not implemented in model | |
| $\frac{1}{\chi}$ | Age interval (years) | 5 | |

### Vaccine supply schedule

Table S6 shows the exemplary vaccine supply schedule used in this analysis. Monthly supply was proportioned into a weekly delivery schedule. Supply numbers are based on publicly reported numbers[21,22] and proportioned to represent a decreasing supply constraint over time. However, these numbers should not be interpreted as actual supply projections for Canada.

*Table S6. Vaccine supply schedule used in analysis*

| **Month** | **Supply** |
|---|---|
| January (4 weeks) | 1.2M |
| February (4 weeks) | 1.8M |
| March (5 weeks) | 3M |
| April (4 weeks) | 3.9M |
| May (5 weeks) | 7.8M |
| June (4 weeks) | 7.8M |
| July (4 weeks) | 7.8M |
| August (5 weeks) | 23.4M |
| September (4 weeks) | 27.3M |
| October (4 weeks) | 0 |
| November (5 weeks) | 0 |
| December (4 weeks) | 0 |

### Supplementary analyses

#### *Probabilistic analyses: $VE_{inf}$ = 40-60% relative to $VE_{dis}$*

A scenario assuming lower effectiveness against infection ($VE_{inf}$ = 40-60% $VE_{dis}$) continued to project reductions in symptomatic disease, hospitalizations, and deaths in the population with extended intervals (Figure S5). The two $VE_{inf}$ scenarios (40-60% and 80-95% relative to $VE_{dis}$) projected very similar results indicating that the benefits of extended intervals were not sensitive to this assumption.

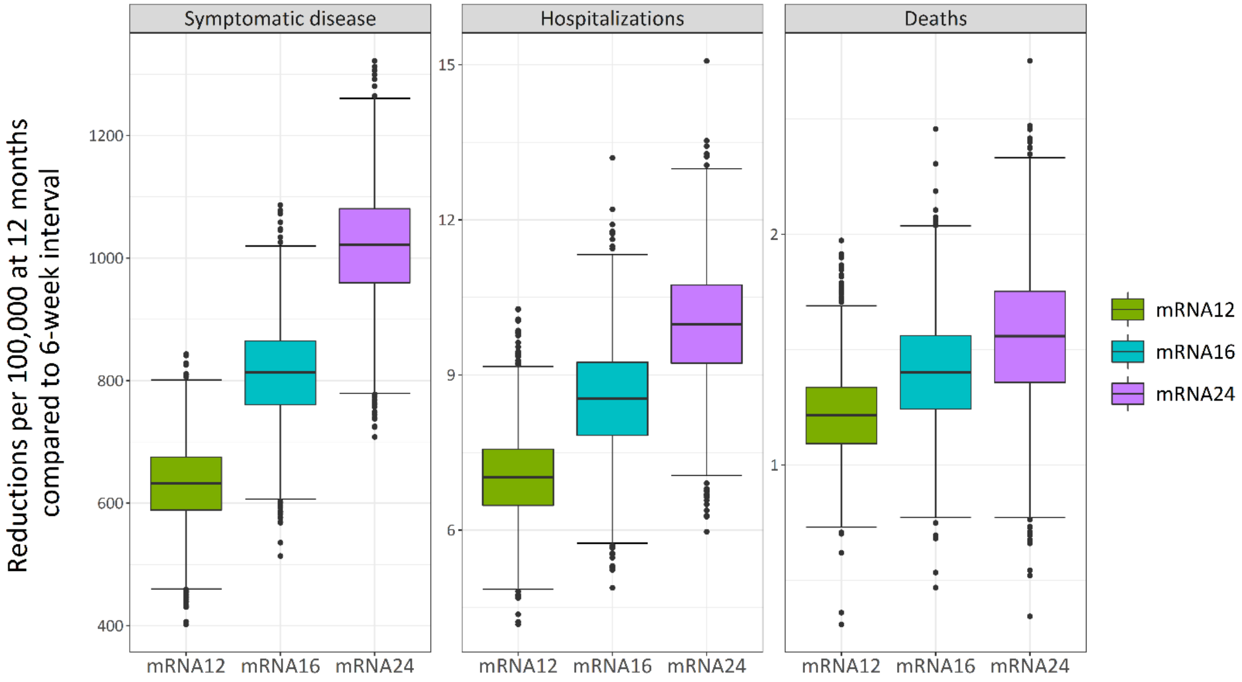


Figure S5. Reductions in symptomatic disease, hospitalizations, and deaths at 12 months compared to a 6-week interval (mRNA6) over 2,000 sampled values of vaccine effectiveness with $VE_{inf}$ = 40-60% $VE_{dis}$.

#### *3$^{rd}$ wave scenarios*

Figure S6 shows how a less severe wave ($R_{eff}$ = 1.1) than the base case ($R_{eff}$ = 1.2) results in an increase in the $VE_{death}$ threshold (from 65% to 70%). In Figure S7, a more severe wave ($R_{eff}$ = 1.3) lowers the $VE_{death}$ threshold from (65% to 60%). If the risk of infection and severe outcomes increases later in the year, a lower first-dose effectiveness against death can still prevent more deaths in exchange for deferred higher protection in high-risk individuals early in the year when the risk is lower. Conversely, if the risk of infection and severe outcomes starts to decrease later in the year (or is lower relative other epidemic scenarios), the relative benefit of accelerating protection in the population waiting for vaccines begins to decrease as there are fewer deaths to prevent, thus requiring a higher first dose effectiveness against death to have greater benefit than shorter intervals.

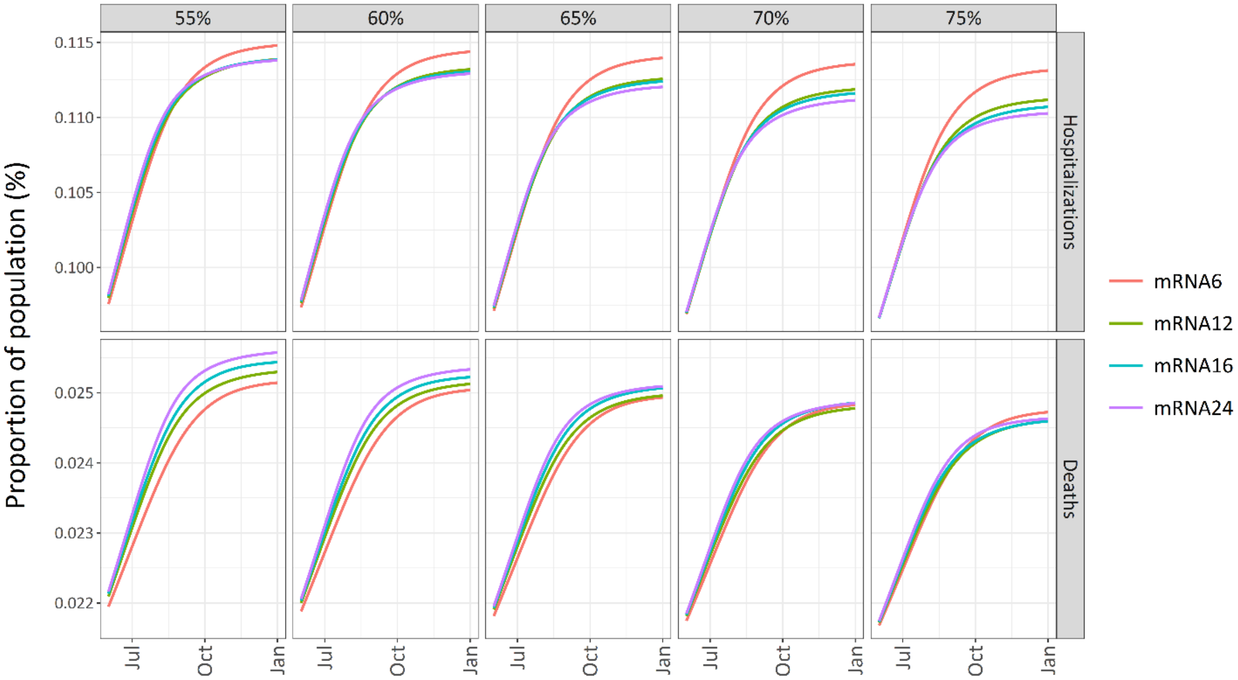


*Figure S6. Cumulative hospitalizations and deaths over $VE_{hosp}$ and $VE_{death}$ values with a 3rd wave beginning on April 1, 2021 ($R_{eff}$ = 1.1 in the absence of vaccinations). $VE_{dis}$ = 50% and $VE_{inf}$ = 90% $VE_{dis}$.*

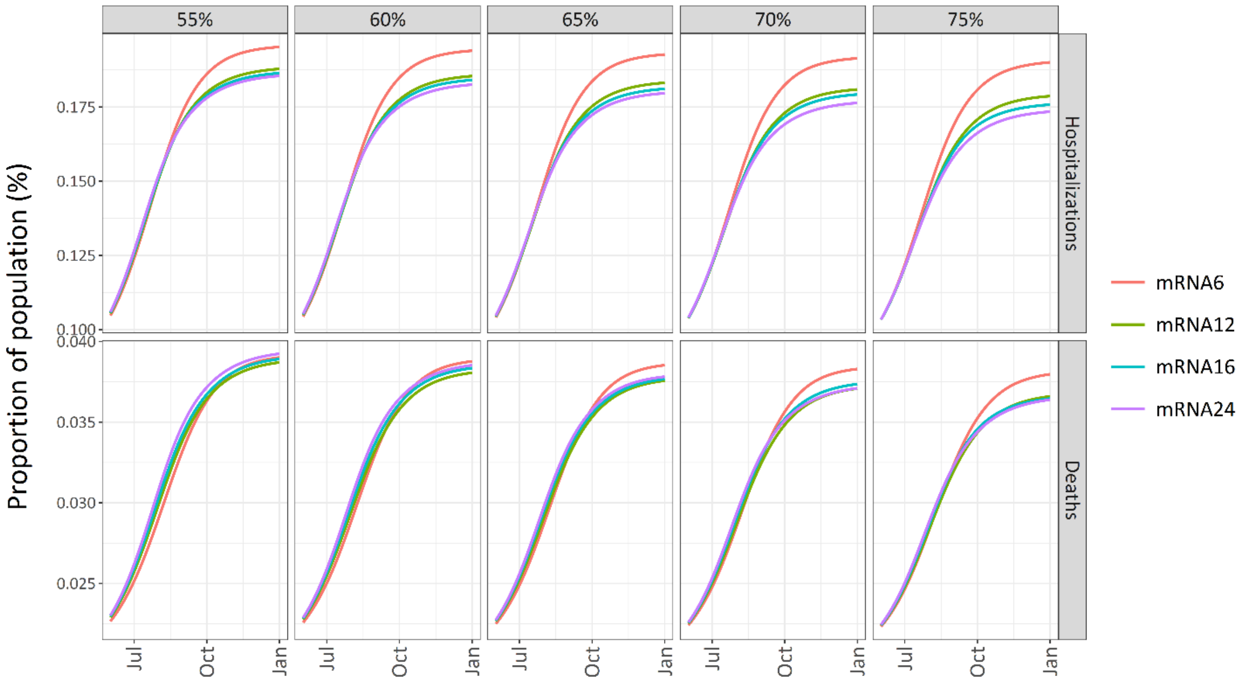


*Figure S7. Cumulative hospitalizations and deaths over $VE_{hosp}$ and $VE_{death}$ values with a 3rd wave beginning on April 1, 2021 ($R_{eff}$ = 1.3 in the absence of vaccinations). $VE_{dis}$ = 50% and $VE_{inf}$ = 90% $VE_{dis}$.*

### References

1. Public Health Ontario. iPHIS Resources [Internet]. 2021 [cited 2021 Jan 11]. Available from: https://www.publichealthontario.ca/en/diseases-and-conditions/infectious-diseases/ccm/iphis

2. Ontario Treasury Board Secretariat. Status of COVID-19 Cases in Ontario [Internet]. 2021.

3. Ontario Agency for Health Protection and Promotion (Public Health Ontario). Enhanced epidemiological summary: COVID-19 in long-term care home residents in Ontario – January 15, 2020 to June 1, 2020. Toronto, ON; 2020.

4. ter Braak CJF, Vrugt JA. A Markov Chain Monte Carlo version of the genetic algorithm Differential Evolution: easy Bayesian computing for real parameter spaces. Stat Comput. 2006;16(3):239–49.

5. van Ravenzwaaij D, Cassey P, Brown SD. A simple introduction to Markov Chain Monte–Carlo sampling. Psychon Bull Rev. 2018;25(1):143–54.

6. Byambasuren O, Cardona M, Bell K, Clark J, McLaws M-L, Glasziou P. Estimating the extent of asymptomatic COVID-19 and its potential for community transmission: Systematic review and meta-analysis. Off J Assoc Med Microbiol Infect Dis Canada. 2020 Dec 1;5(4):223–34.

7. Zhao S. Estimating the time interval between transmission generations when negative values occur in the serial interval data: using COVID-19 as an example. Math Biosci Eng. 2020;17(4):3512–9.

8. He X, Lau EHY, Wu P, Deng X, Wang J, Hao X, et al. Temporal dynamics in viral shedding and transmissibility of COVID-19. Nat Med. 2020;26(5):672–5.

9. Public Health Agency of Canada. Canada COVID-19 Weekly Epidemiology Report 17 January to 23 January 2021 [Internet]. 2021. Available from: https://www.canada.ca/content/dam/phac-aspc/documents/services/diseases/2019-novel-coronavirus-infection/surv-covid19-weekly-epi-update-20210129-eng.pdf

10. Canadian Institute for Health Information. COVID-19 Hospitalization and Emergency Department Statistics, 2019–2020 and 2020–2021. Ottawa,ON: CIHI; 2020.

11. McCarthy Z, Xiao Y, Scarabel F, Tang B, Bragazzi NL, Nah K, et al. Quantifying the shift in social contact patterns in response to non-pharmaceutical interventions. J Math Ind. 2020;10(1):28.

12. Prem K, Cook AR, Jit M. Projecting social contact matrices in 152 countries using contact surveys and demographic data. PLOS Comput Biol. 2017 Sep 12;13(9):e1005697.

13. Heymann AD, Zacay G, Shasha D, Bareket R, Kadim I, Sikron FH, et al. BNT162b2 Vaccine Effectiveness in Preventing Asymptomatic Infection with SARS-CoV-2 Virus: A Nationwide Historical Cohort Study. Available SSRN https//ssrn.com/abstract=3796868 or https//dx.doi.org/102139/ssrn3796868.

14. Shah AS V, Gribben C, Bishop J, Hanlon P, Caldwell D, Wood R, et al. Effect of vaccination on

transmission of COVID-19: an observational study in healthcare workers and their households. medRxiv. 2021 Jan 1;2021.03.11.21253275.

15. Amit S, Regev-Yochay G, Afek A, Kreiss Y, Leshem E. Early rate reductions of SARS-CoV-2 infection and COVID-19 in BNT162b2 vaccine recipients. Lancet. 2021 Mar 6;397(10277):875–7.

16. Pawlowski C, Lenehan P, Puranik A, Agarwal V, Venkatakrishnan AJ, Niesen MJM, et al. FDA-authorized COVID-19 vaccines are effective per real-world evidence synthesized across a multi-state health system. medRxiv. 2021 Jan 1;2021.02.15.21251623.

17. Dagan N, Barda N, Kepten E, Miron O, Perchik S, Katz MA, et al. BNT162b2 mRNA Covid-19 Vaccine in a Nationwide Mass Vaccination Setting. N Engl J Med. 2021 Feb 24;

18. Haas EJ, Angulo FJ, McLaughlin JM, Anis E, Singer SR, Khan F, et al. Nationwide Vaccination Campaign with BNT162b2 in Israel Demonstrates High Vaccine Effectiveness and Marked Declines in Incidence of SARS-CoV-2 Infections and COVID-19 Cases, Hospitalizations, and Deaths. Available SSRN https//ssrn.com/abstract=3811387.

19. Baden LR, El Sahly HM, Essink B, Kotloff K, Frey S, Novak R, et al. Efficacy and Safety of the mRNA-1273 SARS-CoV-2 Vaccine. N Engl J Med. 2020 Dec 30;384:403–16.

20. Polack FP, Thomas SJ, Kitchin N, Absalon J, Gurtman A, Lockhart S, et al. Safety and Efficacy of the BNT162b2 mRNA Covid-19 Vaccine. N Engl J Med. 2020 Dec 10;383(27):2603–15.

21. Tasker JP. Canada to receive 1 million COVID-19 vaccine doses a week starting in April: general [Internet]. CBC News. 2021. Available from: https://www.cbc.ca/news/politics/vaccine-rollout-update-fortin-1.5872766

22. Tasker JP. More Pfizer shots will arrive in 2nd quarter than originally planned: Trudeau [Internet]. CBC News. 2021. Available from: https://www.cbc.ca/news/politics/more-pfizer-shots-trudeau-1.5912209
